# Long-Term Follow-Up Defines the Population That Benefits from Early Interception in a High-Risk Smoldering Multiple Myeloma Clinical Trial Using the Combination of Ixazomib, Lenalidomide, and Dexamethasone

**DOI:** 10.1101/2024.04.19.24306082

**Authors:** Omar Nadeem, Michelle P. Aranha, Robert Redd, Michael Timonian, Sophie Magidson, Elizabeth D. Lightbody, Jean-Baptiste Alberge, Luca Bertamini, Ankit K. Dutta, Habib El-Khoury, Mark Bustoros, Jacob P. Laubach, Giada Bianchi, Elizabeth O’Donnell, Ting Wu, Junko Tsuji, Kenneth Anderson, Gad Getz, Lorenzo Trippa, Paul G. Richardson, Romanos Sklavenitis-Pistofidis, Irene M. Ghobrial

## Abstract

**Background:** Early therapeutic intervention in high-risk SMM (HR-SMM) has demonstrated benefit in previous studies of lenalidomide with or without dexamethasone. Triplets and quadruplet studies have been examined in this same population. However, to date, none of these studies examined the impact of depth of response on long-term outcomes of participants treated with lenalidomide-based therapy, and whether the use of the 20/2/20 model or the addition of genomic alterations can further define the population that would benefit the most from early therapeutic intervention. Here, we present the results of the phase II study of the combination of ixazomib, lenalidomide, and dexamethasone in patients with HR-SMM with long-term follow-up and baseline single-cell tumor and immune sequencing that help refine the population to be treated for early intervention studies.

**Methods:** This is a phase II trial of ixazomib, lenalidomide, and dexamethasone (IRD) in HR-SMM. Patients received 9 cycles of induction therapy with ixazomib 4mg on days 1, 8, and 15; lenalidomide 25mg on days 1-21; and dexamethasone 40mg on days 1, 8, 15, and 22. The induction phase was followed by maintenance with ixazomib 4mg on days 1, 8, and 15; and lenalidomide 15mg d1-21 for 15 cycles for 24 months of treatment. The primary endpoint was progression-free survival after 2 years of therapy. Secondary endpoints included depth of response, biochemical progression, and correlative studies included single-cell RNA sequencing and/or whole-genome sequencing of the tumor and single-cell sequencing of immune cells at baseline.

**Results:** Fifty-five patients, with a median age of 64, were enrolled in the study. The overall response rate was 93%, with 31% of patients achieving a complete response and 45% achieving a very good partial response or better. The most common grade 3 or greater treatment-related hematologic toxicities were neutropenia (16 patients; 29%), leukopenia (10 patients; 18%), lymphocytopenia (8 patients; 15%), and thrombocytopenia (4 patients; 7%). Non-hematologic grade 3 or greater toxicities included hypophosphatemia (7 patients; 13%), rash (5 patients; 9%), and hypokalemia (4 patients; 7%). After a median follow-up of 50 months, the median progression-free survival (PFS) was 48.6 months (95% CI: 39.9 – not reached; NR) and median overall survival has not been reached. Patients achieving VGPR or better had a significantly better progression-free survival (p<0.001) compared to those who did not achieve VGPR (median PFS 58.2 months vs. 31.3 months). Biochemical progression preceded or was concurrent with the development of SLiM-CRAB criteria in eight patients during follow-up, indicating that biochemical progression is a meaningful endpoint that correlates with the development of end-organ damage. High-risk 20/2/20 participants had the worst PFS compared to low- and intermediate-risk participants. The use of whole genome or single-cell sequencing of tumor cells identified high-risk aberrations that were not identified by FISH alone and aided in the identification of participants at risk of progression. scRNA-seq analysis revealed a positive correlation between MHC class I expression and response to proteasome inhibition and at the same time a decreased proportion of GZMB+ T cells within the clonally expanded CD8+ T cell population correlated with suboptimal response.

**Conclusions:** Ixazomib, lenalidomide and dexamethasone in HR-SMM demonstrates significant clinical activity with an overall favorable safety profile. Achievement of VGPR or greater led to significant improvement in time to progression, suggesting that achieving deep response is beneficial in HR-SMM. Biochemical progression correlates with end-organ damage. Patients with high-risk FISH and lack of deep response had poor outcomes. ClinicalTrials.gov identifier: (NCT02916771)

## Introduction

Smoldering multiple myeloma (SMM), defined by the presence of ≥ 10% clonal plasma cells in the bone marrow and/or ≥ 3g/dl monoclonal (M) protein in the serum without the presence of any myeloma-defining events (MDE), carries a heterogenous risk of progression to overt multiple myeloma (MM), with an average risk of 10% per year for the first 5 years after diagnosis and then approximately 1-2% per year thereafter^1^. There is a subset of patients that are considered high-risk SMM (HR-SMM) and multiple efforts have been undertaken to further define this high-risk group, which carries an approximately 50% risk of progression to overt MM at 2 years. The Mayo 2008 criteria defined HR-SMM as having ≥10% plasmacytosis, M protein ≥3 g/dl, and an SFLC ratio ≥ 8 or ≤ 0·125^2^. Many additional risk models have been studied including the PETHEMA criteria, evolving criteria, and IgA subtype, all of which were also shown to have prognostic relevance^3^. A study by Rajkumar et al. compiled all the high-risk features of SMM to help define this population^4^. Later, the IMWG/Mayo 2018 criteria were revised and defined high risk as having the presence of at least 2 of the following 3 criteria: bone marrow plasmacytosis >20%, M protein >2 g/dl, and an SFLC ratio >20^5,6^. While these models help identify patients at high risk of progression, they mostly leverage clinical variables and can give discordant results for an individual^7^. Prediction models that take into account disease biology may be able to overcome some of these challenges. In recent years for example, *MYC* translocations and copy number abnormalities, *KRAS* mutations, Del17p and *TP53* mutations, as well as APOBEC activity and chromothripsis have been shown to portend a higher risk of progression^8–12^.

Management of SMM has historically consisted of observation. However, with advances in risk stratification methods and MM therapy, early therapeutic intervention may offer an improved prognosis without morbidity from MM for high-risk patients. There have been 2 phase III trials utilizing lenalidomide or lenalidomide and dexamethasone in patients with HR-SMM. Both studies demonstrated improvement in progression-free survival (PFS), and one also demonstrated improvement in overall survival, further arguing for early intervention^13,14^. While the prolongation of overall survival with early intervention is a subject of debate, prolonging progression-free survival may delay end-organ damage which can be irreversible and can severely impact a patient’s quality of life^15–17^.

Following these two randomized studies, multiple phase II studies using triplet or more recently quadruplet therapy have been initiated in high-risk SMM^18–21^. However, there are many unresolved questions about the optimal management of patients with HR-SMM, particularly regarding the use of response assessment metrics that were originally developed in patients with overt MM, such as depth of response and biochemical progression, as well as determining the group of patients who may benefit the most from early intervention. The studies using lenalidomide with or without dexamethasone had low rates of complete response and therefore could not assess the role of depth of response in long-term follow-up. Additionally, more recent studies using quadruplet therapy do not have enough long-term follow-up to assess this question yet. Here, we used the combination of ixazomib, lenalidomide, and dexamethasone in high-risk SMM. We aimed to help address several critical questions for therapeutic intervention in HR-SMM: 1) whether depth of response (VGPR or CR) matters in the long-term outcome of patients treated with a lenalidomide-based therapy, 2) whether biochemical progression correlates with end-organ damage, 3) whether 20/2/20 defines participants who would benefit the most from early intervention with lenalidomide-based therapy, and 4) whether the addition of genomic biomarkers can improve the risk stratification of these participants enrolled on these studies.

## Results

### Baseline characteristics

Between March 2017 and February 2020, 55 patients were enrolled on this study (**Figure 1A**). Patient baseline characteristics, definition of high-risk SMM, and high-risk cytogenetics are summarized in **Table 1A**. The median age was 64 (range 40-84) with 30 (55%) males and 25 (45%) females. All patients had an Eastern Cooperative Oncology Group (ECOG) performance status of ≤1. Patients were enrolled using the list of high-risk criteria compiled by Rajkumar et al^4^. For cross-study comparisons, we defined patients with high-risk disease based on several validated models in **Table 1B**, including the Mayo 2008, Mayo 2018, IMWG risk score tool, and high-risk cytogenetics. High risk FISH was defined as follows: gain1q, del13q, del17p, del1p, t(4;14), t(14;16).

**Figure 1.**
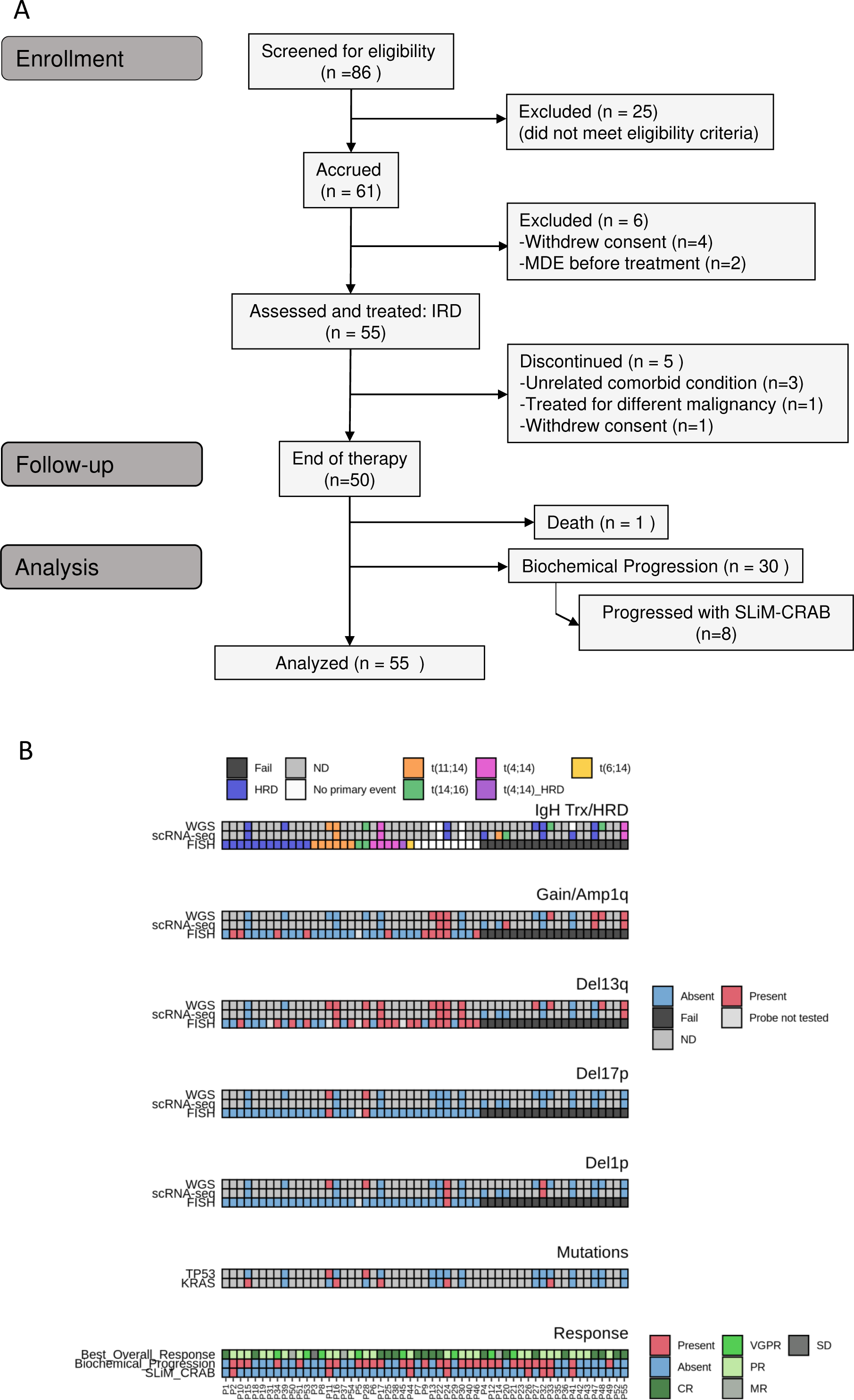
**A)** Consort Diagram **B)** Primary events such as translocations and hyperdiploidy and secondary events such as copy number alterations identified by FISH, WGS, and scRNA-seq along with response to therapy. WGS = Whole genome sequencing, scRNA = Single-cell RNA sequencing. scRNA-seq revealed a more complex aberration in one case (HRD with trisomies of chromosomes 5, 9, 11, 15, 19, and 21) compared to whole-genome sequencing (WGS) which only detected trisomies on chromosomes 9 and 15. This explains the apparent discrepancy between WGS finding “no primary event” and scRNA-seq identifying HRD (homologous recombination deficiency) in that case. ND=Not determined, CR=Complete response, VGPR=Very good partial response, PR=Partial response, MR=Minimal response, SD=Stable disease.

**Table 1A.**
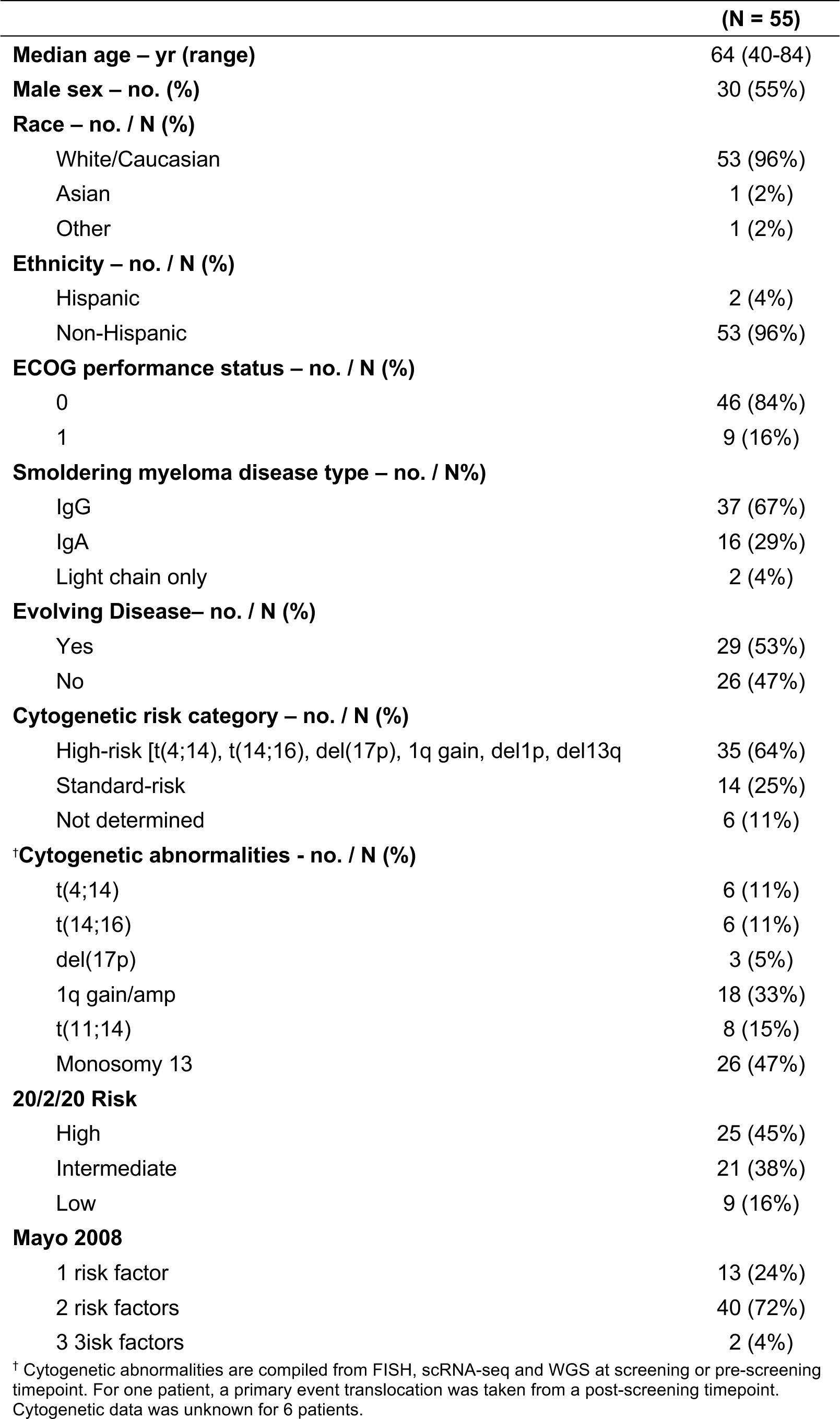
Baseline Characteristics.

**Table 1B.** Baseline Risk Stratification.

| Patient ID | IgA Isotype<sup>a</sup> | M-spike $\geq 3.0^b$ | Immunoparesis<sup>c</sup> | FLC Ratio $\geq 8^d$ | Evolving Disease<sup>e</sup> | BMPC 50-60%<sup>f</sup> | t(4;14)-17p+1q<sup>g</sup> | High-Risk cytogenetics FISH<sup>h</sup> | High-Risk cytogenetics FISH+Seq<sup>i</sup> | Mayo 2008 | IMWG 20/2/20 | IMWG High Precision | Number of High-Risk Criteria † | Total number of eligibility criteria met (+BMPC $\geq 10\%$) †† |
| --- | --- | --- | --- | --- | --- | --- | --- | --- | --- | --- | --- | --- | --- | --- |
| P1 | 1 | 0 | 0 | 1 | 1 | 0 | 0 | 0 | 0 | Int | Int | Low-Int | 1 | 3 |
| P2 | 0 | 1 | 1 | 1 | 1 | 1 | 1 | 1 | 1 | High | High | High | 5 | 6 |
| P3 | 1 | 0 | 1 | 0 | 0 | 0 | 0 | 0 | 0 | Low | Int | Low | 0 | 2 |
| P4 | 0 | 0 | 1 | 1 | 1 | 0 | N/A | N/A | 0 | Int | Low | Low | 1 | 3 |
| P5 | 0 | 0 | 0 | 1 | 1 | 0 | 0 | 1 | 1 | Int | High | High | 4 | 2 |
| P6 | 1 | 0 | 0 | 0 | 0 | 0 | 1 | 1 | 1 | Low | Low | Low | 1 | 2 |
| P7 | 1 | 0 | 1 | 0 | 0 | 0 | 0 | 1 | 1 | Low | Int | Low-Int | 1 | 2 |
| P8 | 1 | 0 | 1 | 0 | 0 | 0 | 0 | 0 | 0 | Low | Low | Low | 0 | 2 |
| P9 | 0 | 0 | 1 | 1 | 0 | 0 | 0 | 0 | 1 | Int | High | Low-Int | 2 | 2 |
| P10 | 0 | 0 | 1 | 1 | 0 | 1 | 1 | 1 | 1 | Int | Int | High | 2 | 4 |
| P11 | 0 | 0 | 1 | 1 | 1 | 0 | 1 | 1 | 1 | Int | Int | Int | 2 | 4 |
| P12 | 1 | 0 | 0 | 0 | 1 | 0 | 1 | 1 | 1 | Low | Low | Low | 2 | 3 |
| P13 | 0 | 0 | 1 | 1 | 0 | 0 | 1 | 1 | 1 | Int | Low | Int | 1 | 3 |
| P14 | 0 | 0 | 1 | 0 | 0 | 0 | N/A | N/A | N/A | Low | High | Low-Int | 1 | 1 |
| P15 | 0 | 1 | 0 | 0 | 1 | 1 | 0 | 0 | 0 | Int | High | Int | 2 | 3 |
| P16 | 0 | 0 | 0 | 1 | 1 | 0 | 0 | 1 | 1 | Int | High | Int | 3 | 2 |
| P17 | 0 | 1 | 0 | 1 | 0 | 1 | 1 | 1 | 1 | High | High | High | 4 | 4 |
| P18 | 0 | 0 | 1 | 1 | 0 | 0 | 0 | 0 | 0 | Int | High | High | 2 | 2 |
| P19 | 1 | 0 | 1 | 1 | 0 | 0 | 0 | 0 | 0 | Int | Int | Int | 0 | 3 |
| P20 | 0 | 0 | 0 | 1 | 0 | 0 | N/A | N/A | 1 | Int | Int | Low-Int | 1 | 1 |
| P21 | 1 | 0 | 0 | 0 | 1 | 0 | N/A | N/A | N/A | Low | Low | Low | 1 | 2 |
| P22 | 0 | 0 | 0 | 1 | 0 | 0 | 1 | 1 | 1 | Int | High | High | 3 | 2 |
| P23 | 0 | 0 | 0 | 1 | 1 | 0 | N/A | N/A | N/A | Int | Low | Low | 1 | 2 |
| P24 | 0 | 0 | 1 | 1 | 1 | 1 | 1 | 1 | 1 | Int | High | High | 4 | 5 |
| P25 | 1 | 0 | 1 | 1 | 0 | 1 | 1 | 1 | 1 | Int | High | Int | 2 | 5 |
| P26 | 0 | 0 | 0 | 0 | 0 | 0 | 1 | 1 | 1 | Low | Low | Low-Int | 1 | 1 |
| P27 | 0 | 0 | 0 | 1 | 0 | 0 | 0 | 1 | 1 | Int | Int | Low-Int | 1 | 1 |
| P28 | 0 | 0 | 1 | 0 | 1 | 0 | 1 | 1 | 1 | Low | High | Int | 3 | 3 |
| P29 | 0 | 0 | 0 | 1 | 1 | 0 | 0 | 0 | 0 | Int | Int | Low-Int | 1 | 2 |
| P30 | 1 | 0 | 1 | 1 | 1 | 1 | 0 | 1 | 1 | Int | High | Int | 3 | 5 |
| P31 | 0 | 0 | 1 | 1 | 1 | 0 | 0 | 0 | 0 | Int | High | Low-Int | 2 | 3 |
| P32 | 0 | 0 | 1 | 1 | 1 | 1 | 0 | 0 | 1 | Int | High | High | 4 | 4 |
| P33 | 0 | 0 | 1 | 1 | 1 | 1 | 1 | 1 | 1 | Int | Int | Low-Int | 2 | 5 |
| P34 | 0 | 0 | 0 | 1 | 0 | 0 | 1 | 1 | 1 | Int | High | Int | 2 | 2 |
| P35 | 0 | 0 | 0 | 1 | 1 | 0 | N/A | N/A | N/A | Int | Int | Low-Int | 1 | 2 |
| P36 | 0 | 0 | 1 | 1 | 0 | 0 | 1 | 1 | 1 | Int | Int | Int | 1 | 3 |
| P37 | 1 | 0 | 1 | 1 | 1 | 0 | 0 | 0 | 0 | Int | Int | Int | 1 | 4 |
| P38 | 0 | 0 | 1 | 1 | 1 | 0 | 1 | 1 | 1 | Int | High | High | 4 | 4 |
| P39 | 0 | 0 | 1 | 0 | 0 | 0 | 0 | 0 | 0 | Low | High | Low-Int | 1 | 1 |
| P40 | 0 | 0 | 1 | 1 | 1 | 0 | 0 | 1 | 1 | Int | Low | Low-Int | 2 | 3 |
| P41 | 0 | 0 | 0 | 1 | 0 | 0 | N/A | N/A | 0 | Int | Int | Low-Int | 0 | 1 |
| P42 | 1 | 0 | 1 | 1 | 1 | 0 | 0 | 1 | 1 | Int | Int | Low-Int | 2 | 4 |
| P43 | 0 | 0 | 0 | 1 | 1 | 0 | N/A | N/A | N/A | Int | Int | Low-Int | 1 | 2 |
| P44 | 0 | 0 | 1 | 1 | 1 | 0 | 0 | 1 | 1 | Int | Int | Int | 2 | 3 |
| P45 | 1 | 0 | 1 | 1 | 1 | 0 | 1 | 1 | 1 | Int | Int | Low-Int | 2 | 5 |
| P46 | 1 | 0 | 1 | 1 | 1 | 0 | 1 | 1 | 1 | Int | High | Int | 3 | 5 |
| P47 | 1 | 0 | 0 | 0 | 0 | 0 | 1 | 1 | 1 | Low | Int | Low | 1 | 2 |
| P48 | 0 | 0 | 1 | 0 | 0 | 0 | 0 | 1 | 1 | Low | Int | Low-Int | 1 | 1 |
| P49 | 0 | 0 | 1 | 1 | 1 | 0 | N/A | N/A | N/A | Int | High | Low-Int | 2 | 3 |
| P50 | 0 | 0 | 0 | 1 | 1 | 0 | 0 | 1 | 1 | Int | High | Int | 3 | 2 |
| P51 | 0 | 0 | 1 | 0 | 0 | 0 | 0 | 0 | 0 | Low | High | Low-Int | 1 | 1 |
| P52 | 1 | 0 | 1 | 1 | 0 | 0 | N/A | N/A | N/A | Int | High | Int | 1 | 3 |
| P53 | 1 | 0 | 0 | 1 | 0 | 0 | 1 | 1 | 1 | Int | Int | Low-Int | 1 | 3 |
| P54 | 1 | 0 | 0 | 1 | 0 | 0 | 0 | 1 | 1 | Int | High | Low-Int | 2 | 2 |
| P55 | 0 | 0 | 0 | 1 | 1 | 0 | N/A | N/A | 1 | Int | High | High | 4 | 2 |

Abbreviations: FLC, Free Light Chain; BMPC, Bone Marrow Plasma Cells; FISH, Fluorescent In-Situ Hybridization; Seq, Sequencing; Int, Intermediate.
Columns <sup>a-g</sup> contain high-risk criteria as defined by Rajkumar et al<sup>4</sup>. High-Risk cytogenetics in columns <sup>h,i</sup> defined as t(4;14), t(14;16), 1q gain/amp, 13q deletion/monosomy 13, dedeletion 1p, and 17p deletion.
† Total Number of High-Risk Criteria by Mayo 2008, IMWG 20/2/20, IMWG High Precision, Evolving Disease, and High-Risk Cytogenetics by FISH+Seq
†† Total Eligibility criteria met for enrollment as described in Methods section.

While FISH testing is a standard method to define risk in MM, the failure rate of FISH in this study was 36% (FISH at screening was successful in 35/55) (**Figure 1B**). Therefore, we employed whole genome sequencing (WGS) and single-cell RNA (scRNA) sequencing to identify high-risk translocations and copy number alterations (CNA). The translocations and copy number alterations identified by FISH, WGS, and scRNA-seq are displayed in **Figure 1B**. From scRNA-seq, we leveraged the expression level of primary IgH translocation marker genes to infer the presence of translocations and observed high concordance between the expression of said marker genes and FISH results, but additionally detected translocations in three patients in whom FISH had failed due to low cell numbers. Furthermore, using Numbat^22^ to infer CNAs from scRNA-seq data, we detected hyperdiploidy (homologous recombination deficiency: HRD) in another four patients in whom FISH had failed due to low cell numbers, as well as additional high-risk secondary CNVs such as Amp1q, Del13q, and Del1p in four patients. Whole-genome sequencing confirmed the IgH translocations, hyperdiploidy, and secondary CNVs in 10 patients with both WGS and scRNA-seq of tumor cells. No discrepancies were observed between scRNA-seq and WGS. Overall, for ten patients where FISH failed due to a low number of cells, scRNA-seq and WGS data existed and we were able to resolve the cytogenetic abnormalities for these cases. Notably, the first aspirate pull was used for clinical-grade FISH testing, while later pulls were used for research-level WGS and scRNA-seq; given the frequent hemodilution observed in later aspirate pulls which further decreases the number of tumor cells in the specimen, these results showcase the superior sensitivity in tumor cell detection and characterization of these methods compared to FISH. Further, by reviewing pre-screening FISH reports and incorporating primary translocation data from treatment records for remaining failures, we obtained cytogenetic data for 49 out of 55 individuals. Since we did not have whole-exome sequencing or WGS data on all patients, we refrained from assessing the impact on outcomes of certain high-risk mutations, including MAPK mutations, DNA repair mutations, or MYC alterations, which have been previously identified as high risk^8,10,12^.

### Safety

Toxicity results are summarized in **Table 2**. Thirty-six percent of patients experienced a grade 2 toxicity, 62% experienced a grade 3 toxicity and 9% experienced a grade 4 toxicity. The most common treatment-related toxicities of any grade included leukopenia in 44 patients (80%), neutropenia in 43 patients (78%), fatigue in 42 patients (76%), rash in 38 patients (69%), and diarrhea in 37 patients (67%). The most common grade 3 treatment-related hematologic toxicities were neutropenia in 14 patients (25%), leukopenia in 10 patients (18%), lymphopenia in 6 patients (11%), and thrombocytopenia in 4 patients (7%). Non-hematologic grade 3 toxicities included hypophosphatemia in 7 patients (13%) and rash in 5 patients (9%). Grade 4 toxicities related to study treatment included hyperglycemia (n = 1/55; 2%), hypokalemia (n = 1/55; 2%), lymphopenia (n = 2/55; 4%), neutropenia (n = 2/55; 4%), and thrombocytopenia (n = 1/55; 2%). One patient developed a grade 5 intracranial hemorrhage 3 weeks after completion of therapy that was not related to the study intervention; the patient was on therapeutic anticoagulation with a history of atrial fibrillation and the event was considered unrelated by the patient’s treating physicians.

**Table 2.**
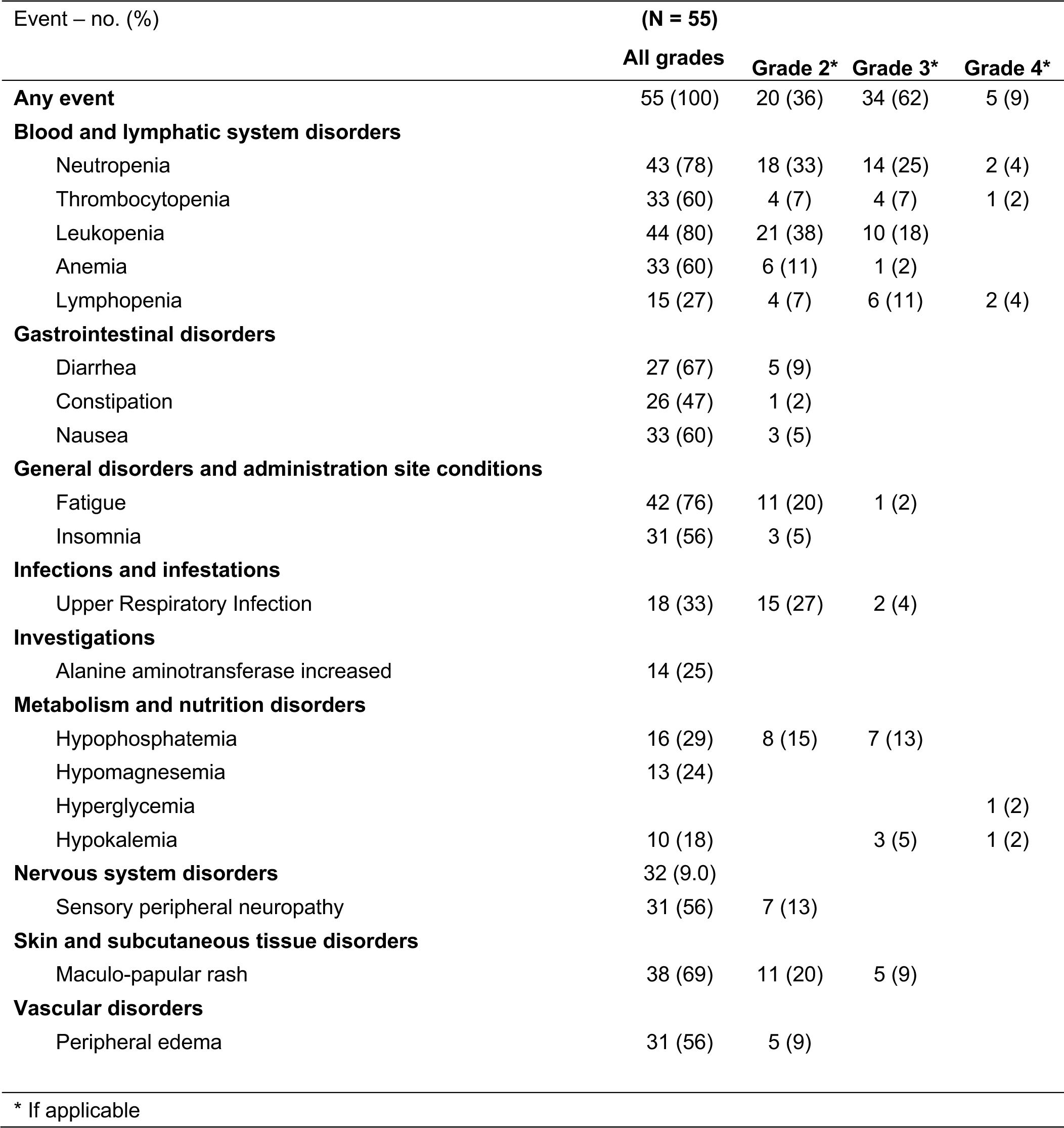
Summary of Adverse Reactions Reported During Treatment.

| Event – no. (%) | (N = 55) |  |  |  |
| --- | --- | --- | --- | --- |
|  | All grades | Grade 2* | Grade 3* | Grade 4* |
| **Any event** | 55 (100) | 20 (36) | 34 (62) | 5 (9) |
| **Blood and lymphatic system disorders** |  |  |  |  |
| Neutropenia | 43 (78) | 18 (33) | 14 (25) | 2 (4) |
| Thrombocytopenia | 33 (60) | 4 (7) | 4 (7) | 1 (2) |
| Leukopenia | 44 (80) | 21 (38) | 10 (18) |  |
| Anemia | 33 (60) | 6 (11) | 1 (2) |  |
| Lymphopenia | 15 (27) | 4 (7) | 6 (11) | 2 (4) |
| **Gastrointestinal disorders** |  |  |  |  |
| Diarrhea | 27 (67) | 5 (9) |  |  |
| Constipation | 26 (47) | 1 (2) |  |  |
| Nausea | 33 (60) | 3 (5) |  |  |
| **General disorders and administration site conditions** |  |  |  |  |
| Fatigue | 42 (76) | 11 (20) | 1 (2) |  |
| Insomnia | 31 (56) | 3 (5) |  |  |
| **Infections and infestations** |  |  |  |  |
| Upper Respiratory Infection | 18 (33) | 15 (27) | 2 (4) |  |
| **Investigations** |  |  |  |  |
| Alanine aminotransferase increased | 14 (25) |  |  |  |
| **Metabolism and nutrition disorders** |  |  |  |  |
| Hypophosphatemia | 16 (29) | 8 (15) | 7 (13) |  |
| Hypomagnesemia | 13 (24) |  |  |  |
| Hyperglycemia |  |  |  | 1 (2) |
| Hypokalemia | 10 (18) |  | 3 (5) | 1 (2) |
| **Nervous system disorders** | 32 (9.0) |  |  |  |
| Sensory peripheral neuropathy | 31 (56) | 7 (13) |  |  |
| **Skin and subcutaneous tissue disorders** |  |  |  |  |
| Maculo-papular rash | 38 (69) | 11 (20) | 5 (9) |  |
| **Vascular disorders** |  |  |  |  |
| Peripheral edema | 31 (56) | 5 (9) |  |  |

\* If applicable

Five patients developed secondary malignancies (one instance each of prostate, head/neck, ovarian, uterine and melanoma). Three of five patients were diagnosed following the completion of treatment on protocol and remained on follow-up while receiving active treatment for secondary malignancy. The remaining two patients were diagnosed during active treatment; one participant delayed treatment for secondary malignancy (prostate) to finish protocol therapy and remained on study through the end of protocol-based follow-up. The other patient diagnosed during active treatment discontinued therapy to receive treatment for their secondary malignancy (melanoma) and remained on follow-up. No patients were diagnosed with myelodysplastic syndrome or other myeloid malignancies.

Dose modifications occurred in 34 patients (62%) during the study therapy. The median number of modifications was 1 (range 1-4) and dose delays due to toxicity occurred in 19 patients (35%). The most common reason for dose modification of ixazomib was peripheral neuropathy (10 patients, 18%). Stem cells were collected from all eligible patients by the end of the induction phase and there was one patient who failed to mobilize stem cells.

The median number of cycles completed was 24 (range: 2-24). Five patients discontinued therapy prior to the planned 24 cycles. Reasons for discontinuation included worsening unrelated co-morbid conditions (n=3), treatment for a different malignancy (n=1), and withdrawal of consent (n=1). No patients discontinued treatment due to toxicity.

### Efficacy

The overall response rate (partial response or better) based on International Myeloma Working Group (IMWG) criteria^23^ was 93% (n=51), with complete response (CR) observed in 17 patients (31%), VGPR in 8 patients (15%), and partial response in 26 patients (47%) (**Table 3**). All patients who achieved CR also achieved a stringent and sustained CR for at least 6 months with responses deepening over time **(Figure 2)**. Twenty-five patients (45%) achieved a VGPR or better response and minimal response or better was observed in 98% of patients (n=54). The overall response rate was similar for patients in different 20/2/20 risk groups: low (100%), intermediate (90%), and high (92%) (Cochran-Armitage test, p=0.55).

**Figure 2.**
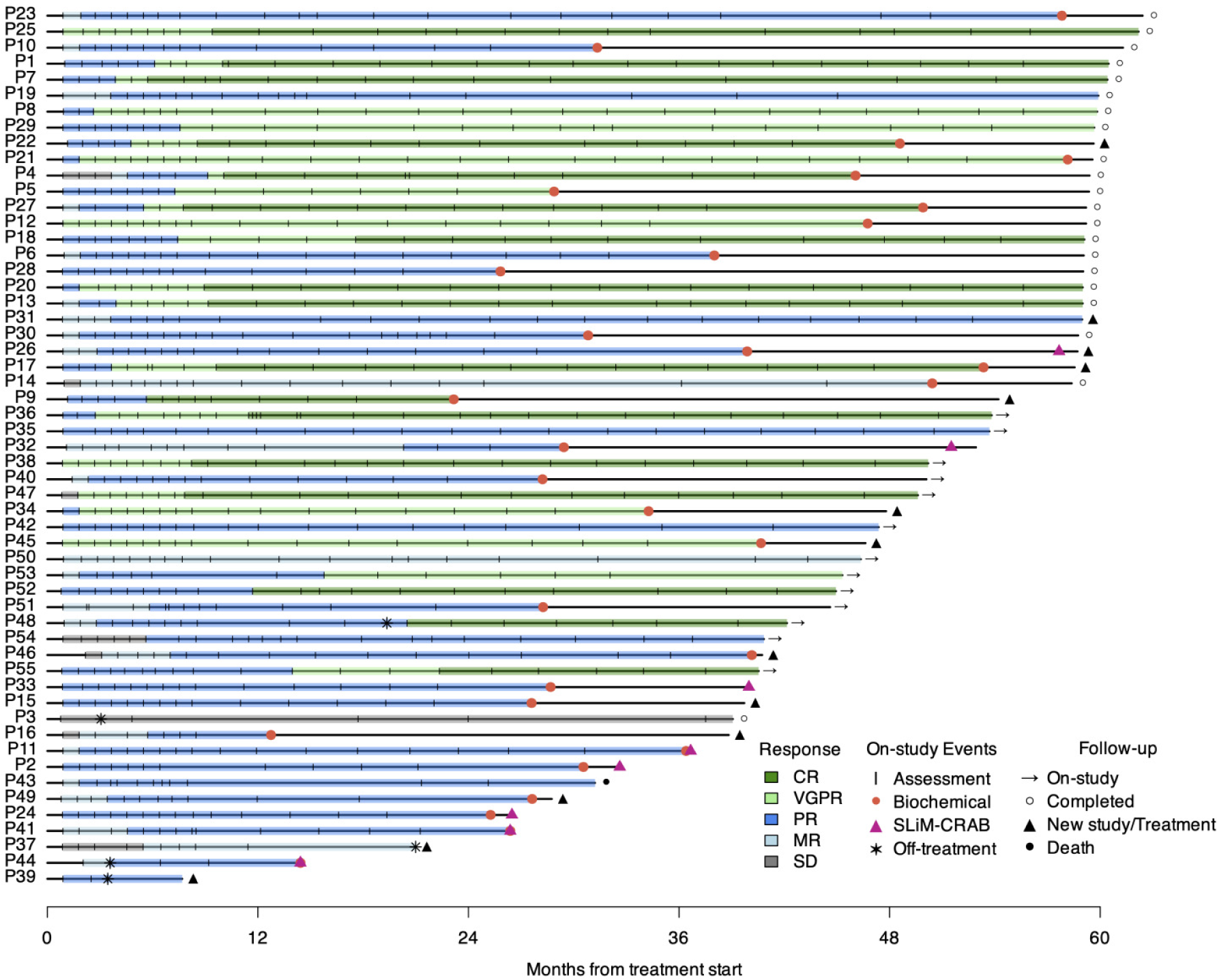
Swimmer’s plot of responses deepening over time. Each lane represents one participant.

**Table 3.**
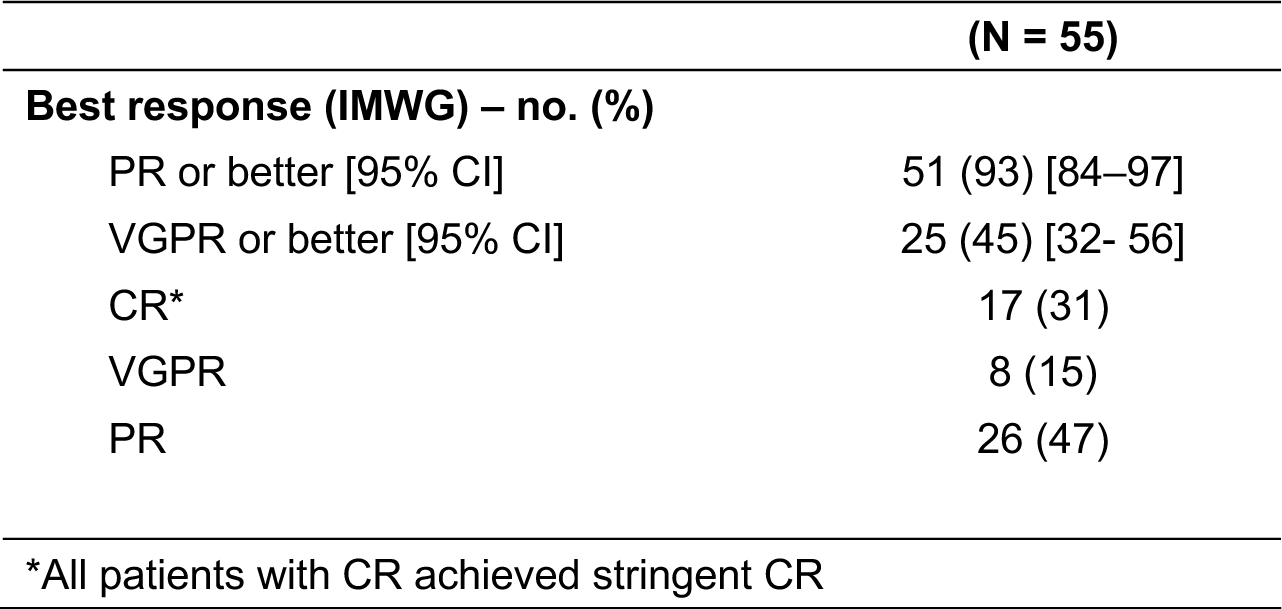
Summary of Responses to Treatment.

Eight participants progressed to overt MM defined by the SLiM-CRAB myeloma-defining events (MDEs)^24^ (**Figure 2**). All eight patients experienced biochemical progression concurrent with or prior to developing SLiM-CRAB progression (**Figure 2**).One patient discontinued protocol therapy after one cycle due to unrelated comorbid conditions including colitis and atrial fibrillation and remained on follow-up for ten months prior to SLiM-CRAB progression. The remaining seven progressors completed all twenty-four cycles of protocol therapy and continued on follow-up prior to progression. Four of eight progressors had an MDE within one year of their final treatment, and the median time to progression for the remaining four progressors was 21 months (range: 13.2-32.7 months). Four patients developed lytic bone disease noted on PET and MRI during follow-up, one patient developed renal failure and hypercalcemia 9 months after completion of therapy (after showing biochemical progression at 6 months post-therapy), one patient developed anemia 17 months post-therapy after showing biochemical progression at 6 months post-therapy, one patient met SLiM-CRAB criteria at 13 months post-therapy with FLC ratio > 100 and BMPCs of 60% prompting initiation of therapy and one patient met SLiM-CRAB criteria at 13 months post-therapy with FLC ratio >100. Seven of the eight had at least one high-risk genetic event at baseline [1q gain (n=4), Del13q (n=5), Del17p (n=2, one confirmed from FISH tests and one identified in a pre-screening FISH report from patient records), Del1p (n=3), t(14;16) (n=1), KRAS (n=2), TP53 (n=1)] (**Figure 1B**). Among all progressors, the best response achieved was PR with none achieving VGPR or better.

### Time-to-event analyses

The median follow-up for all 55 patients was 50 months (range: 8-61 months). Thirty patients had biochemical progression events (**Figure 2**). Biochemical progression was defined as a 25% increase in serum or urine M protein or a difference between involved and uninvolved FLC levels based on the IMWG criteria^25^.

The total number of patients who were followed for at least two years and remained biochemical progression-free (including the censored patients who came off therapy with or without progression) was 50 of 55 patients (91%; 90% CI: 82 - 96%, p < 0.001, one-sided exact binomial test) **(Figure 3A)**.

**Figure 3.**
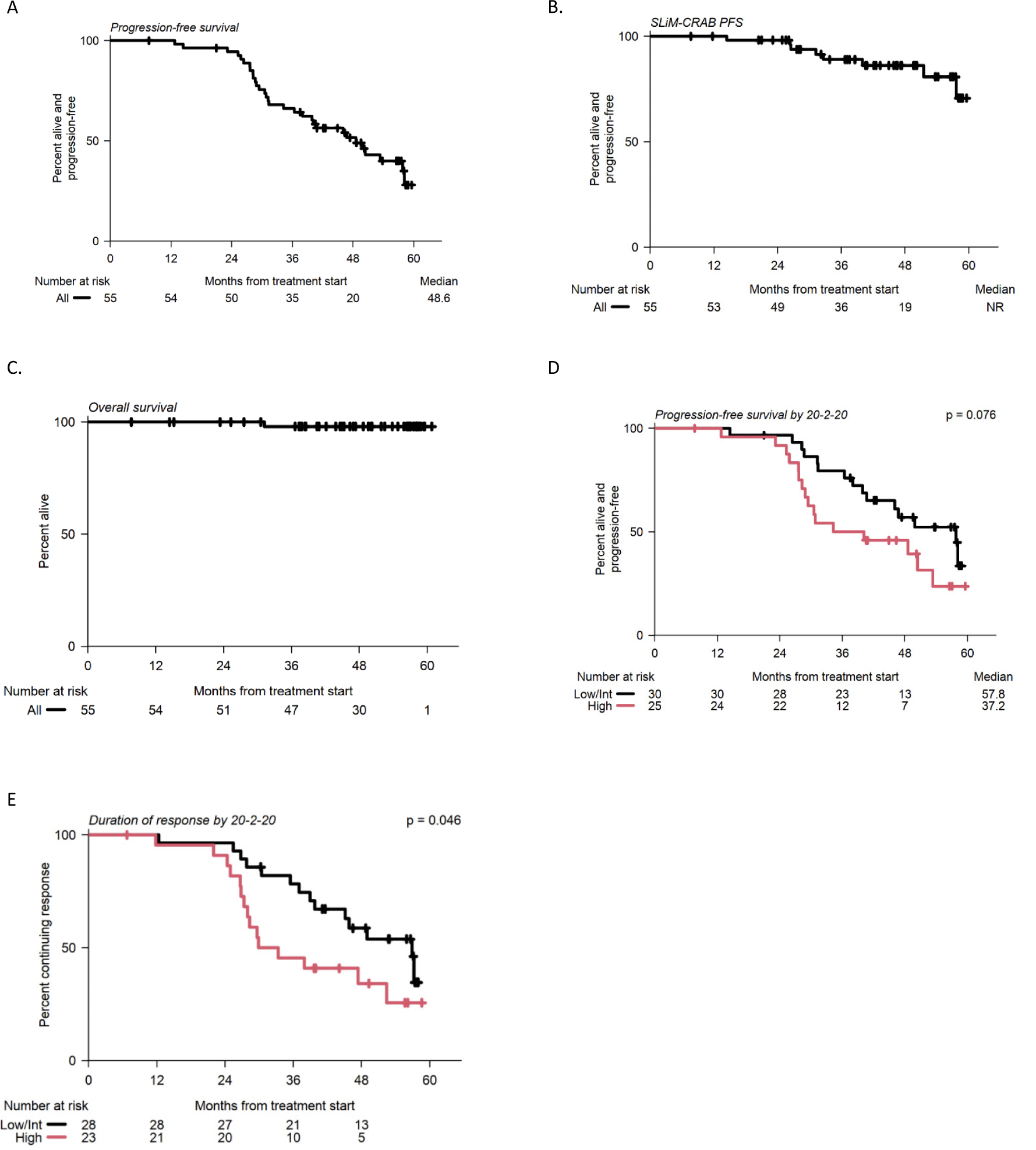
Progression-free and overall survival. Progression defined by **A)** Biochemical progression or **B)** SLiM CRAB criteria **C**) Overall survival. NR = not reached **D**) Kaplan-Meier curve of response duration stratified by 20/2/20 risk groups. **E**) Kaplan-Meier curve of progression stratified by 20/2/20 risk groups. Survival distributions were compared using one-sided log-rank tests.

Progression-free survival for the primary endpoint was defined as the development of end-organ damage per the SLiM-CRAB criteria or death. Participants who were not followed for at least two years, yet remained alive and progression-free while on trial, were censored at the last date known progression-free. The total number of patients who remained progression-free without SLiM-CRAB criteria or death within two years of enrollment was 52 of 53 patients (98%) **(Figure 3B)**. One patient developed SLiM-CRAB progression within 2 years of enrollment after discontinuing therapy (described above). No patients developed SLiM-CRAB progression while on active therapy (**Figure 3B**).

Median biochemical PFS was 48.6 months (95% CI: 39.9 – not reached; NR) and median overall survival has not been reached **(Figure 3 A, C)**. The median duration of response was 47.4 months (95% CI: 37 – NR) and the median time to progression was 49.9 months (95% CI: 39.9 – NR).

### Subsequent treatments for those with biochemical progression but not SLiM-CRAB

Eleven patients started new treatments on SMM-directed clinical trials following the completion of protocol therapy prior to the development of SLiM-CRAB progression. Of these, ten experienced biochemical progression while on follow-up (**Figure 2**) and had an evolving pattern leading to enrollment in a subsequent clinical trial. Evolving pattern was defined as an increase of 10% in the M-spike concentration or the involved free light chain concentration during the 6 months prior to screening for the clinical trial. Four patients met high-risk criteria per the 20/2/20 model on follow-up. These patients elected not to wait for end-organ damage on follow-up and proceeded with therapy on another trial of HR-SMM.

### Factors that impact progression-free survival

We next assessed several baseline characteristics to determine factors that define the population that benefits the most from a lenalidomide-based regimen and further identified endpoints that can be early biomarkers for end-organ damage in this population. We examined baseline characteristics including the 20/2/20 model, evolving subtype, high-risk cytogenetics, the best response to therapy, as well as the combination of response, high-risk cytogenetics, and minimal residual disease (MRD). These criteria were used based on their association with prognosis in prior studies of overt MM in a prior post hoc assessment in lenalidomide studies of SMM^26,27^.

We first determined whether the baseline 20/2/20 criteria helped identify patients who would have a prolonged PFS in response to therapy. We combined low- and intermediate--risk patients and compared them to high-risk patients. Here, we showed that the median PFS was longer but not statistically significant at 57.8 months for the low-/intermediate-risk group compared to 37.2 months for high-risk patients (log-rank, p = 0.08) (**Figure 3D**). The median duration of response was significantly shorter at 31.6 months in high-risk patients compared to 56.9 in the low/int group (log-rank, p = 0.046) **(Figure 3E)**.

Evolving disease was assessed in all participants as part of the eligibility criteria and as described above this was defined as a 10% or greater increase in either the M-spike concentration or the involved free light chain concentration within the 6 months preceding clinical trial screening. Of all patients that enrolled in the clinical trial (N=55), an evolving pattern was found in 29 out of 55 patients (53%) while 26 out of the 55 patients (47%) did not have an evolving pattern (**Table 1**). The median time of the lab result was 107 days prior to the screening date with an interquartile range of 120 days. The presence of evolving disease at baseline was similar across baseline IMWG 20/2/20 risk groups: low (56%), intermediate (48%), and high (56%) (Cochran-Armitage, p=0.83). Of the patients that demonstrated an evolving pattern, only 31% (n=9/29) achieved a VGPR or better, and of those that did not demonstrate an evolving pattern, 62% (n=16/26) achieved a VGPR or better (Fisher’s exact, p=0.032).

In patients who did not demonstrate an evolving pattern, the PFS was significantly longer at 53.4 months compared to 40.1 months in those who had evolving disease prior to registration (log-rank, p=0.025). Additionally, in patients who did not demonstrate an evolving pattern, the median duration of response was not reached and was significantly longer compared to that in patients who had an evolving subtype (37.9 months; log-rank, p=0.034) **(Figure 4A-D)**.

**Figure 4.**
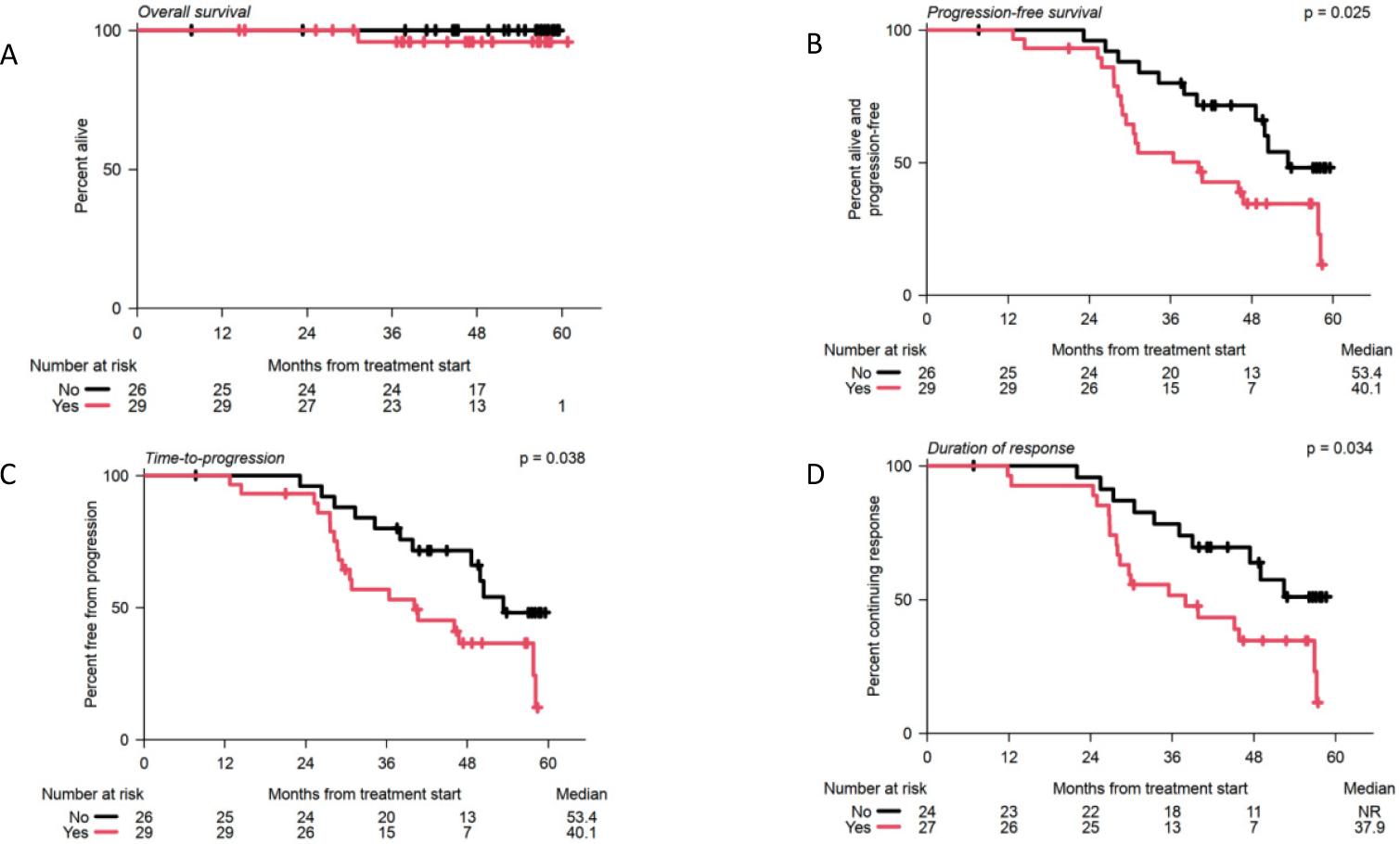
Outcomes stratified by the presence of evolving subtype prior to treatment. **A)** Overall survival **B)** progression-free survival **C)** time-to-progression and **D)** duration of response. NR = not reached. Survival distributions were compared using one-sided log-rank tests.

We next examined whether patients with VGPR or CR had an improved progression-free survival. Response has been associated with PFS in studies of overt MM^28^. However, this has not been previously shown in studies of smoldering myeloma partially because the previous studies using lenalidomide and dexamethasone had few cases of CR and partially for the short follow-up period of the quadruplet regimens being currently assessed in SMM. We, therefore, aimed to determine whether depth of response is associated with progression in the SMM setting. Indeed, we demonstrated that patients who showed VGPR or better had a median time to progression (TTP) of 58.2 months compared to 31.3 months in patients who showed inferior responses (log rank, p < 0.001; **Figure 5 A)**. Interestingly, this was still true in patients with high-risk cytogenetics as determined by FISH only (median PFS 31.3 months vs 53.4 months, log-rank, p = 0.002) or by FISH and whole genome sequencing(WGS) (median PFS of 30.8 months vs 53.4 months, p < 0.001), demonstrating that depth of response is relevant for prognostication independent of cytogenetics (**Figure 5 B,C**).

**Figure 5.**
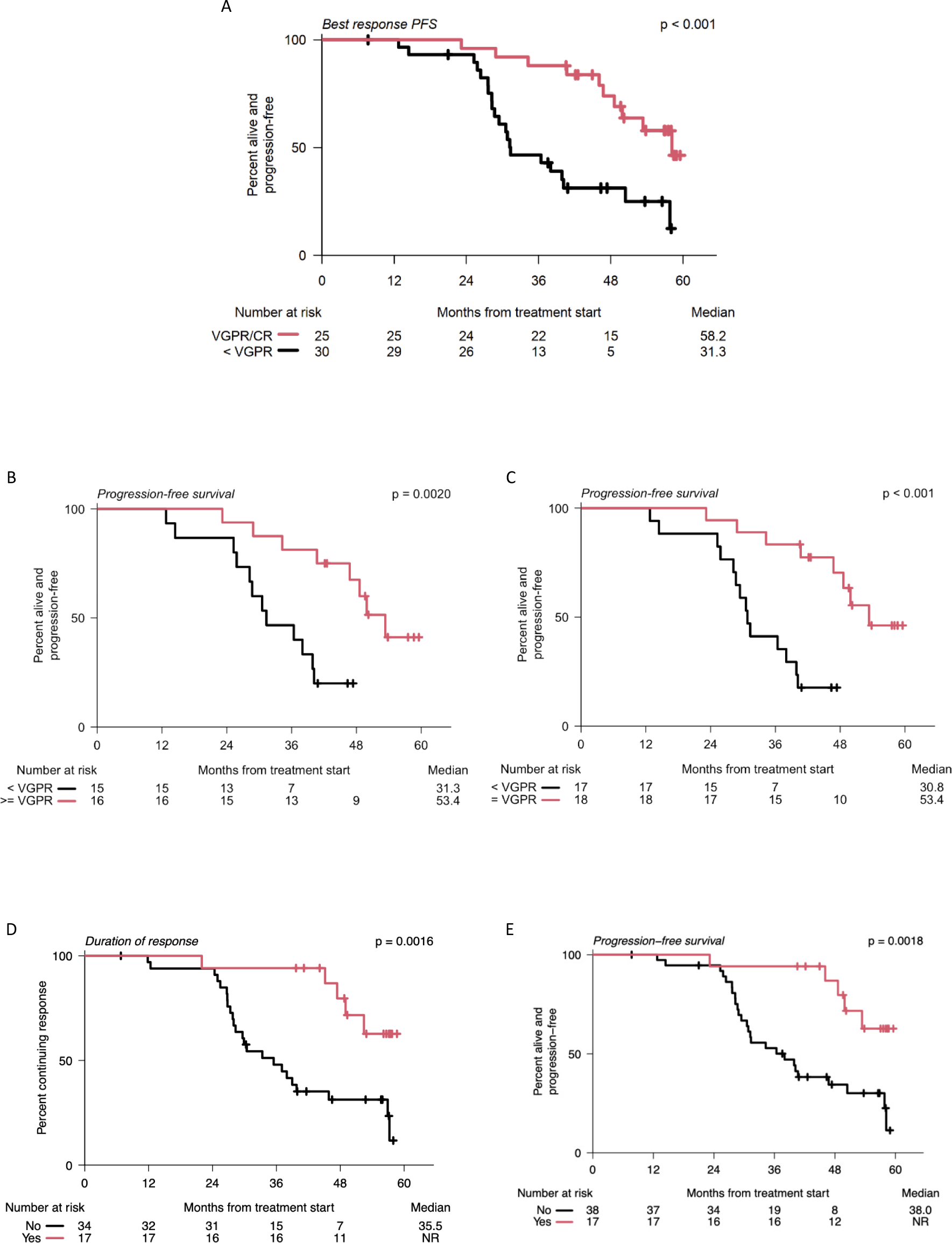
Kaplan Meier Curve of Progression-Free Survival based on. **A**) Depth of Response of VGPR of greater. VGPR = Very good partial response **B**) depth of response amongst cytogenetically high-risk patients by High-risk patients identified by FISH alone. **C**) by High-risk patients identified by FISH, WGS, or single-cell RNA sequencing. **D**) Kaplan-Meier curve of duration of response stratified by patients who achieved sustained CR. **E**) Kaplan-Meier curve of progression-free-survival stratified by patients who achieved sustained CR. CR = Complete response. Survival distributions were compared using one-sided log-rank tests.

All patients who achieved a complete response (CR) maintained it for at least 6 months. The median duration of response in the sustained CR group was not reached at the time of analysis, while the median duration of response in the non-CR group was 35.5 months (log-rank, p = 0.0016) (**Figure 5D**). Similarly, the median progression-free survival in the sustained CR group was not reached, compared to 38 months in the non-CR group (log-rank, p = 0.0018) (**Figure 5E**).

### Minimal Residual Disease

Twenty-five patients were evaluable for MRD (achieved VGPR or better) by next-generation sequencing (NGS), 11 out of 25 had no samples available for MRD testing or had a quality control (QC) failure. Twenty-three samples at C9 or EOT from 14 out of 25 patients passed the QC analysis and were available for MRD testing. A sensitivity of 10^−5^ was reached in 22/23 (96%) samples and a sensitivity of 10^−6^ in 17/23 samples (74%) (**Suppl Figure S1).** Out of patients who showed a VGPR or better response and had MRD testing (n=14), 29% (4/14) were MRD negative at 10^−5^ at EOT and none developed biochemical or clinical progression over the follow-up period. Of those who were MRD positive at 10^−5^ (n=10/14), 6 patients developed biochemical progression during the follow-up period. Two MRD time points (C9 and EOT) were available in a subgroup of 8 individuals. Of those, 50% (4/8) were MRD positive at 10^−5^ at C9 and remained so at EOT, while 50% (4/8) were MRD negative at 10^−5^ at C9 but only 1 patient remained MRD negative at EOT, and 3 converted to MRD positive. The 3 patients that converted from a C9 MRD negative at a sensitivity of 10^−5^ to positive at EOT developed biochemical progression during the follow-up period. The 5-year progression-free survival rate of patients who achieved MRD negativity at 10^−5^ was 100%, compared to 40% of MRD-positive patients (log-rank, p = 0.051) (**Figure 6**).

**Figure 6.**
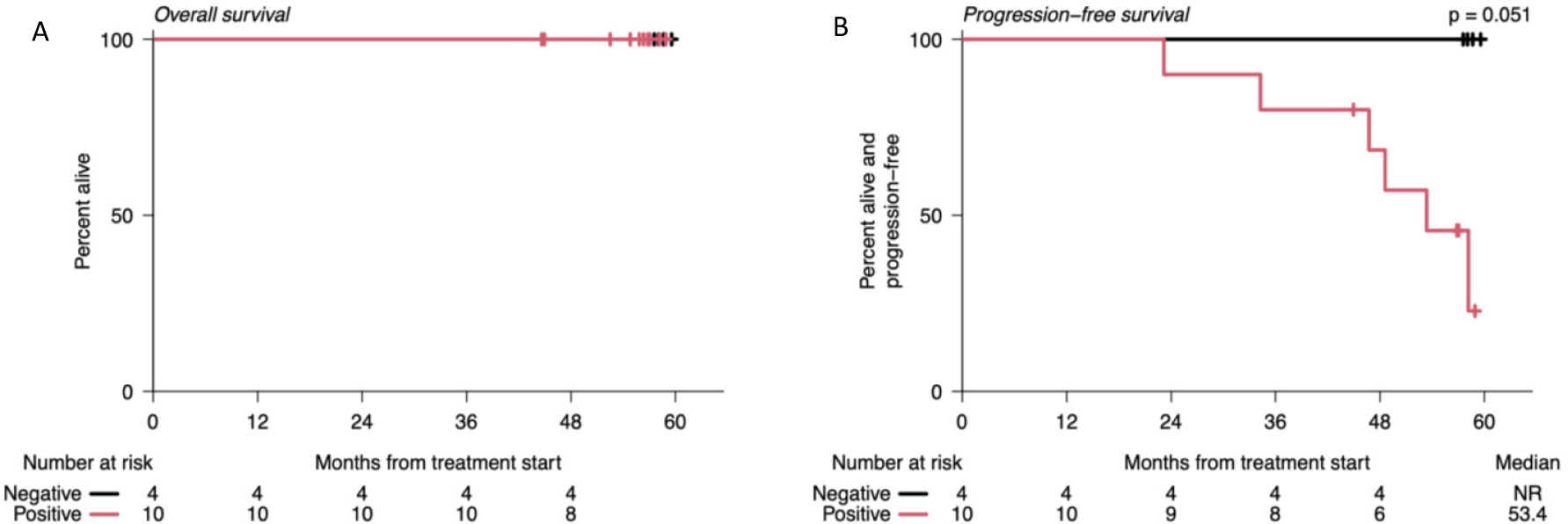
Kaplan Meier Curve of Outcomes Based on MRD Status. **A).** Overall Survival, **B**) Progression-free survival. Survival distributions were compared using one-sided log-rank tests.

### Mass Spectrometry MRD assessment

MALDI-TOF mass spectrometry was available at baseline and C9 in 43 patients. The M-protein was undetectable in 4 patients at C9 (10%). Since clearance of M-protein could take more time than tumor cell clearance in the BM, we tested the serum of patients by MALDI-TOF in available serum samples during maintenance at 2-6 months after C9 (at C11 to 16). 37 were retested during maintenance: six were negative (including the 4 that were negative at C9) and 31 remained positive. At EOT, MALDI-TOF was performed for 41 patients. Eight (20%) were negative: 6 out of 8 confirmed a previous negative result, one case turned negative from a previous positive result, and one had a negative result from baseline with no testing during treatment. The concentration calculated using the EXENTi software positively correlated with that calculated via SPEP at diagnosis, C9 and EOT (p<0.0001) (**Suppl Figure S3**). Of the patients with negative MALDI-TOF at EOT (n=8), 4 developed biochemical progression, and none developed SLiM-CRAB progression. In patients who achieved a negative MALDI-TOF result at EOT, the median progression-free survival was 53.46 months compared to 38.01 months in patients who had a positive MALDI-TOF result at EOT (log-rank, p=0.08) (**Suppl Figure S4**).

We compared residual disease detection using serum immunofixation (IFX) with MALDI-TOF in 86 measurements. While both tests are significantly concordant (Table S1, Cohen’s κ=0.64, p<0.001, 95% CI [0.44, 0.84]) and no patients had a positive IFX test and a negative MALDI-TOF test, MALDI-TOF identified some cases missed by IFX (10 IFX-negative MALDI-TOF-positive cases). Therefore, MALDI-TOF might be more sensitive than IFX for detecting residual disease.

### Single-cell RNA sequencing reveals tumor-intrinsic and extrinsic factors associated with disease progression

To identify biological predictors of response and resistance to IxaRD in baseline samples from patients with HR-SMM, we performed single-cell RNA sequencing on CD138+ and CD138-cells as well as WGS on 22 patients enrolled on the study (14 biochemical progressors, 8 non-progressors) and 11 Healthy donors **(Figure 7A)**. Among the progressors in this subcohort, *5* patients met SLiM CRAB criteria **(Figure 7A)**.

**Figure 7.**
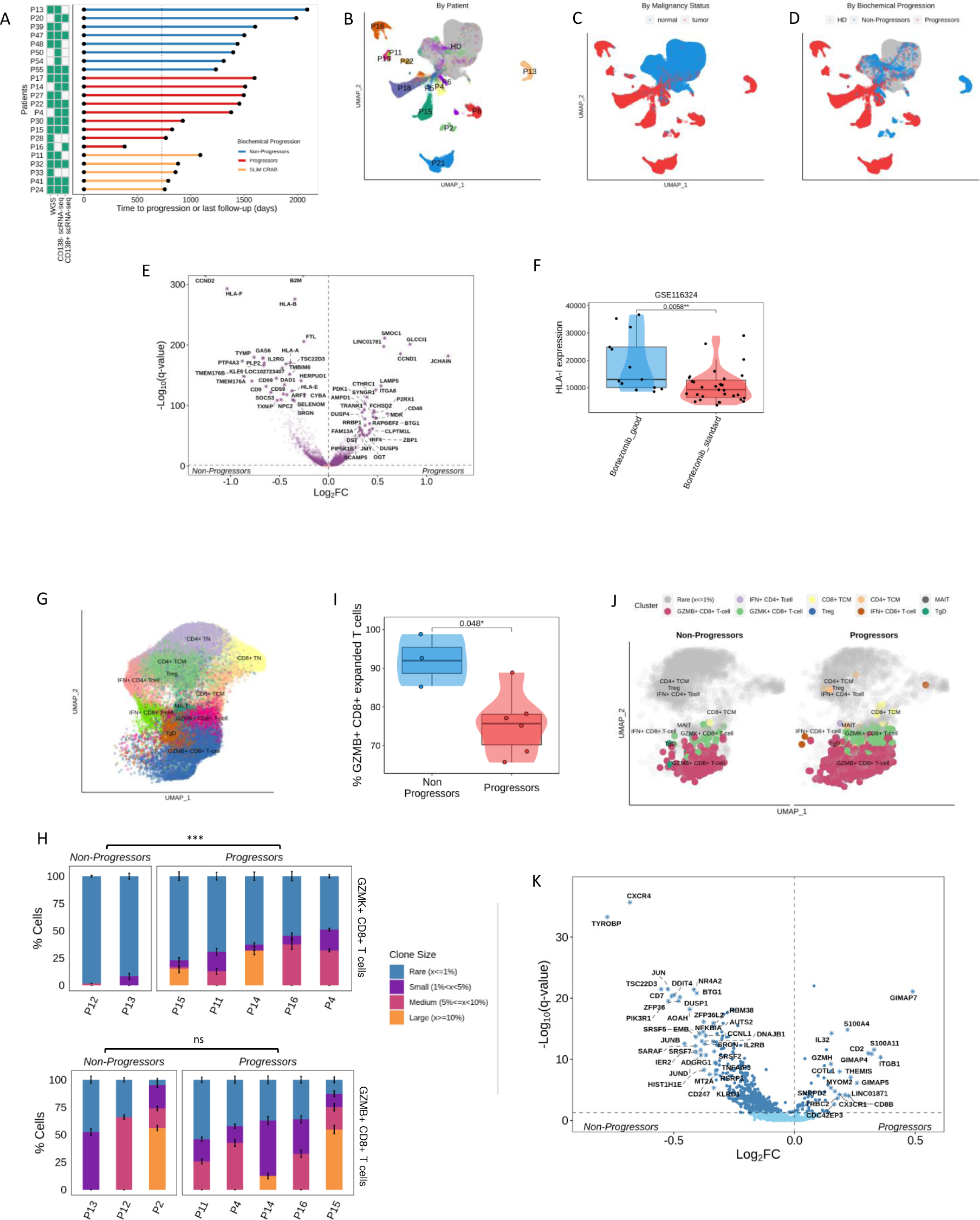
Single-cell RNA sequencing of bone marrow CD138+ cells. **A)** Time to progression or last follow-up for the subcohort of patients with sequencing data (one patient per row). The type of sequencing data available for individual patients is indicated by green boxes. Patients are grouped by progression status. **B)** Uniform manifold approximation and projection (UMAP) embeddings of patients’ and HD’s plasma cells that passed quality filtering Cells are colored by the patient from which they originate or patients or HD as a group **C)** UMAP colored by malignancy status (tumor vs normal cells). **D)** UMAP colored by biochemical progression status. **E)** Volcano plot of differential gene expression in progressors and non-progressors. Two-sided p values were computed with Wilcoxon’s rank-sum test and corrected using the Benjamini-Hochberg approach. The top 30 genes on either side of the volcano with q < 0.05 are indicated by stars. **F**) HLA class Iexpression in patients from the PADIMAC cohort who responded (bortezomib_good) or did not respond (Bortezomib_standard) to bortezomib treatment.Single-cell RNA sequencing of T cells from CD138-samples. **G**) UMAP of Tcells **H**) Bar plot visualizing the proportion of clonotypes belonging to each of four clone size categories per patient in BM CD138-samples drawn at BL. For patient’s T cell subtype being analyzed, 100 cells were randomly sampled 100 times from non-progressor and progressor patients, and the proportion of expanded (1-rare) clonotypes was compared between the two using bootstrapping with 1000 iterations. The average proportion per clone size category is visualized, and the SD across iterations is depicted in solid-line error bars. **I**) Boxplots, violin plots, and scatterplots comparing the abundance of GZMB+ CD8+ T effector memory (TEM) cells in expanded clones between non-progressors and progressors. Two-sided p values were computed with Wilcoxon’s rank-sum test and corrected using the Benjamini-Hochberg approach. **J**) UMAP embedding of progressor and non-progressor patients’ BM T cells at BL with matched TCR data. T cells with expanded clones (with a frequency > 1%) are colored by cell type. Cells with rare clonotypes are shown in grey. **K**) Volcano plot of differential gene expression of clonally expanded CD8 TEM in progressors and non-progressors. p values were computed with Wilcoxon’s rank-sum test and corrected using the Benjamini-Hochberg approach. Top 30 genes on either side of the volcano with q < 0.05 are indicated by stars. HD = healthy donors; P = patient; WGS = whole-genome sequencing; HRD = hyperdiploidy

Overall, we identified 125,572 plasma cells(Patients: malignant, n=50,591; normal, n=1876; HD: n=73,105) **(Figure 7B-D)**. To identify tumor-intrinsic transcriptomic alterations associated with biochemical progression post-IxaRD in patients with HRSMM, we performed differential expression analysis comparing tumor cells from patients who progressed (n=10) to those who did not (n=3). To restrict the influence of individual tumors on the analysis, as well as to control for the lower number of non-progressors in this sub-cohort, we randomly sampled the same number of tumor cells from progressors and non-progressors (∼2100 per group) and balanced the contribution of each individual towards the group (progressors: ∼210 cells per patient; non-progressors: 700 cells per patient; **Figure 7E)**. Additionally, to control for differences in sequencing depth, we downsampled 20K reads per cell across all cells used in this analysis. We observed consistently higher expression levels of MHC class I genes (*HLA-A, HLA-B, HLA-F, HLA-E, B2M*) in tumor cells from non-progressors to progressors **(Figure 7E)**. To validate this observation, we analyzed bulk RNA-sequencing data from an external cohort of MM patients treated with a Bortezomib-based regimen, a different proteasome inhibitor (PADIMAC, GSE116234). This analysis confirmed that tumor cells from patients who respond well to proteasome inhibition show higher levels of MHC-I genes **(Figure 7F)**. Notably, the PADIMAC study did not use Lenalidomide in combination with proteasome inhibition, which was the case in our study. This supports the notion that the observed association with MHC-I levels is related to proteasome inhibition specifically rather than a particular combination.

These results suggest that antigen presentation via MHC-I by tumor cells may be important for response to proteasome inhibition and implicate the immune system in mediating response. While the immune system has been thought to potentiate the effects of proteasome inhibition through increased neoantigen presentation by immune cells following tumor cell killing, it has not been previously linked to tumor-intrinsic transcriptomic alterations^29^. To pursue this hypothesis further, we turned our attention to the cytotoxic CD8+ T cell compartment, which is tasked with responding to MHC-I-bound tumor-derived antigens. Overall, we sequenced 51,326 T-cells, including 5,045 granzyme K (GZMK)-expressing and 9,026 granzyme B (GZMB)-expressing cytotoxic T cells **(Figure 7G)**. First, we systematically compared the frequency of clonal expansion within the GZMK+ and the GZMB+ compartment between progressors and non-progressors. We observed that the GZMK+ compartment was significantly more clonally expanded in progressors compared to non-progressors (bootstrapping, two-sided p<0.001) **(Figure 7H)**, but this was not seen in the GZMB+ compartment. By comparing the frequency of the GZMB+ phenotype within clonally expanded T cells from progressors and non-progressors, we observed a significant decrease in differentiation towards the GZMB+ phenotype in progressors (Wilcoxon, two-sided p=0.048) **(Figure 7 I, J)**. This suggests that clonally expanded effector cytotoxic T cells from progressors may be less mature compared to non-progressors. To gain more insight into the functionality of clonally expanded GZMB+ cytotoxic T cells, we performed differential expression analysis between progressors and non-progressors. We observed that clonally expanded GZMB+ cytotoxic T cells from non-progressors showed higher expression levels of genes related to cytotoxicity and activation, such as *TYROBP*, *CXCR4*, and members of the AP-1 pathway, compared to progressors **(Figure 7K)**. Clonally expanded GZMB+ cytotoxic T cells from progressors showed higher levels of GIMAP genes, which may be associated with increased apoptosis^30^, and *THEMIS*, which can suppress cytotoxic responses^31^, however higher levels of maturation markers *ITGB1* and *CX3CR1* were also observed, suggesting that the functionality of clonally expanded GZMB+ cytotoxic T cells may be more nuanced.

Overall, this suggests that clonally expanded cytotoxic T cells from progressors are phenotypically less mature, and potentially less functional, compared to those from non-progressors, which could be related to the observed differences in MHC-I expression in tumor cells. This data implicates immune dysfunction as a potential correlate of suboptimal response to proteasome inhibition.

## Discussion

Optimal management of HR-SMM continues to evolve with many novel approaches currently under investigation to deliver highly effective therapy to prevent end-organ damage while limiting toxicity. There have been phase III studies of lenalidomide and dexamethasone demonstrating improvement in progression-free and overall survival and there are ongoing studies utilizing triplets or quadruplets that include monoclonal antibodies with proteasome inhibitors, lenalidomide, and dexamethasone. The ixazomib, lenalidomide, and dexamethasone combination in this study demonstrated high response rates with many patients experiencing deep responses, with VGPR or better in nearly half of the patients treated as well as the benefit of an all-oral approach. No patients progressed to overt MM during study therapy and the regimen was overall well-tolerated. The response rates and safety were comparable to results seen in clinical trials of ixazomib, lenalidomide, and dexamethasone of newly diagnosed and relapsed MM^32,33^. The median PFS of 48.9 months also compares favorably to previously reported clinical trials of triplet combination regimens in SMM^19,20^.

Because of the long follow-up of a median of 50 months and the depth of response achieved in half of the patients, we have an opportunity to answer critical questions about the management of early intervention in HR-SMM.

A controversial question in SMM trials is whether depth of response matters in trials of HR-SMM. Previous studies using lenalidomide alone or lenalidomide and dexamethasone demonstrated a long-term benefit and delay of end organ damage even if the best response was only PR. The argument was that immune equilibrium may be re-instated in those patients and deep remission is not required. However, other studies using “curative intent” argued that achieving a depth of response similar to overt MM is the most optimal way to achieve long-term remission. Here, we show for the first time that a depth of response of VGPR or greater correlated with time to progression, suggesting that deep response is impactful in SMM similar to overt MM. This was particularly highlighted in patients with cytogenetically high-risk disease, where PFS was only 30.8 months in patients with less than VGPR.

The next question was whether MRD is associated with improved PFS. Although the numbers were small, we were still able to see near-significant differences in the PFS of MRD negative cases compared to those who did not achieve or sustain MRD negativity. Ongoing studies with deeper remissions with quadruplet therapy or immunotherapy will help further address this question more definitively.

We then examined the correlation of biochemical progression with the development of SLiM-CRAB criteria. Thirty patients developed biochemical progression in all (27 after completion of therapy), of which 8 patients also developed SLiM-CRAB with overt MM. Eleven patients started new treatments on SMM-directed trials following the completion of protocol therapy prior to developing SLiM-CRAB progression. Of these, ten experienced biochemical progression while on follow-up and had an evolving pattern leading to enrollment in a subsequent clinical trial. Four of these patients met high-risk criteria per the 20/2/20 model. This is consistent with our clinical observation that patients who opt to be treated initially for HR-SMM do not want to wait for the development of end-organ damage prior to re-initiating therapy, consistent with recent discussions to consider biochemical progression as an end-point of therapy rather than SLiM-CRAB^34^. In addition, it brings up a critical question of whether re-treatment on a trial of SMM should be in the same cohort with other participants who were never exposed to therapy or in a separate cohort. Additional guidance is necessary to determine which patients would be suitable for further SMM-directed therapy vs MM induction therapy.

The strong correlation between biochemical progression and MDE also points out that biochemical progression can be used as a surrogate for MDE in high-risk SMM. However, the number of patients is small, and other studies need to further confirm this observation.

Interestingly, the use of 20/2/20 criteria in our study demonstrated that responses were similar across all risk criteria, but the progression-free survival was shortest for those who were high-risk per 20/2/20. This indicates that high-risk 20/2/20 patients may resemble MM and may require more aggressive therapy or maintenance therapy. We did not have an observation control arm in this study, so we cannot assess whether achieving response in these patients truly alters their long-term risk of progression to MM.

Moreover, the use of evolving subtype helped capture those with increasing biomarkers regardless of their disease burden. The best response to treatment was not similar in those who showed increasing biomarkers prior to receiving therapy compared to those who had fairly stable biomarkers. In patients who did not show an evolving disease pattern as defined in our eligibility criteria, the best response to therapy achieved was better, duration of response was longer, and their progression-free survival was superior compared to those who demonstrated increasing biomarkers prior to therapy. Our study highlights the benefit of including evolving disease as an indication of high-risk disease. Future studies will help shed light on the importance of varying biomarkers in terms of risk stratification and the severity of disease.

Genomic alterations are critical to better define patients who are truly at risk of progression and who may benefit from more aggressive therapy. In this study, the addition of WGS and single-cell RNA sequencing helped identify participants with high-risk cytogenetics who otherwise would not have been identified by FISH alone. We opted not to include MAPK and DNA repair pathway mutations in analyzing impact on outcomes because we did not have data available on all patients. With additional data, we were able to indeed define a population of high-risk cytogenetics and poor response to therapy who showed the worst outcome.

Minimal residual disease was performed with two approaches in this protocol: standard FDA-approved NGS on BM samples achieving at least VGPR and very sensitive MALDI-TOF mass spectrometry. MALDI-TOF was assessed on peripheral blood serum, avoiding bone marrow biopsy, and is gaining more and more interest in the field. We showed that MALDI-TOF is a sensitive and accurate assay for monitoring M protein isotype and quantification, with a concordance with SPEP at baseline and a strong prognostic effect after EOT. We showed that MALDI-TOF is superior to IFX with 100% NPV and 83% PPV, similar to previous studies^35^.

We performed an integrative study combining WGS and single-cell RNA-sequencing of tumor and immune cells with the IRD study. To identify tumor-specific gene expression differences that associated with IRD treatment response, we compared gene expression in tumor cells at baseline from progressor and non-progressor patients who received IRD. We observed significantly higher levels of MHC class I genes in tumor cells from non-progressor patients compared to progressors. We further validated this finding using data from an external study on MM patients treated with a different proteasome inhibitor (Bortezomib), suggesting a broader link between MHC-I levels and response to proteasome inhibition. To further explore the immune system’s involvement, we analyzed cytotoxic CD8+ T cells, which recognize and eliminate cells presenting specific antigens on their surface via MHC-I molecule. We compared the clonal expansion of two T cell subsets, granzyme K (GZMK)+ and granzyme B (GZMB)+, and found that progressors exhibited greater clonal expansion in the GZMK+ compartment compared to non-progressors, suggesting a potentially ineffective immune response. We have previously shown that the GZMK+ compartment is the major source of expression of PD-1, a key T cell exhaustion marker ^20^. Additionally, progressors showed a reduced proportion of the GZMB+ phenotype within their clonally expanded T cells, suggesting their clonally expanded T cells have a less mature cytotoxic profile compared to those of non-progressors. We next analyzed functional differences in T cells between progressors and non-progressors. Gene expression patterns revealed higher expression of genes related to cytotoxicity and activation in the non-progressors’ clonally expanded GZMB+ T cells compared to those of the progressors. Clonally expanded GZMB+ T cells from progressors showed increased expression of genes associated with apoptosis and suppression of cytotoxic responses, alongside markers of maturation. This suggests a more complex picture of functionality in these T cells from progressors. Our findings suggest that immune dysfunction related to changes in tumor intrinsic antigen presentation at baseline may contribute to suboptimal responses in proteasome inhibitor therapy.

Our study has several limitations including the non-randomized nature of the trial without a control arm and the potential selection bias in any single-center experience. Several questions remain, including optimal timing of therapy in HR-SMM, duration of therapy, treatment modality (combinations, immunotherapy), and response adaptive approaches. The limited size of the scRNAseq sub-cohort and lack of functional validation limit the conclusiveness of the findings and require further validation in larger, independent cohorts.

In conclusion, this is the first trial studying an all-oral triplet combination of ixazomib, lenalidomide, and dexamethasone in HR-SMM, demonstrating substantial efficacy and identifying several factors critical in the outcome of patients: depth of response, evolving pattern, MRD negativity and high risk 20/2/20. Future studies using quadruplet therapy or immunotherapy may alter these conclusions.

## Methods

The research study was approved by the Dana-Farber/Harvard Cancer Center institutional review board (protocol number DFCI 16-313) and complied with all relevant ethical and legal regulations. All participants gave written informed consent. The clinicaltrials.gov registration number is NCT02916771. Individual deidentified participant sequencing and FISH data will be shared.

### Eligibility Criteria

Patients enrolled on the study met eligibility for high-risk SMM based on criteria by Rajkumar^24^ as follows: bone marrow plasmacytosis ≥ 10% and ≤60% and any one or more of the following: serum M protein ≥3 g/dL, IgA isotype, immune paresis with reduction of 2 uninvolved immunoglobulins, serum involved/uninvolved ratio of ≥8 but ≤100, bone marrow plasmacytosis 50-60%, abnormal plasma cell immunophenotype, high-risk FISH defined as t(4;14), del 17p or 1q gain, increased circulating plasma cells, MRI with diffuse abnormalities or 1 focal lesion, PET/CT with one focal lesion with increased uptake without underlying osteolytic bone destruction and urine monoclonal light chain excretion ≥500mg/24 hours. Patients with evidence of SLiM-CRAB criteria were excluded (bone marrow plasmacytosis > 60%, light chain ratio >100)^36^. Patients were required to have an ECOG performance status of ≤2 and adequate hematologic and organ function prior to enrollment. A creatinine clearance of ≥30 mL/min was required.

### Treatment Regimen

Patients were treated on an outpatient basis with 9 cycles of induction therapy consisting of ixazomib 4mg given orally on days 1, 8, and 15, in combination with lenalidomide 25mg administered orally on days 1 through 21 and dexamethasone 40mg administered orally on days 1, 8, 15, and 22 of a 28-day cycle. The induction phase was followed by maintenance therapy, consisting of ixazomib 4mg on days 1, 8, and 15 and lenalidomide 15mg on days 1-21 without dexamethasone for another 15 cycles. Treatment duration was a total of 24 cycles (2 years).

### Objectives and End Points

The primary objective was to determine the proportion of patients with HR-SMM who are progression-free 2 years after receiving trial therapy. Progression for the primary endpoint was defined as the development of end organ damage per SLiM-CRAB criteria. Secondary objectives included response rates, duration of response, progression-free survival, safety of the combination therapy, and minimal residual disease (MRD) negativity. Mass spectrometry analysis was also performed to correlate with IMWG response and MRD status.

### Minimal Residual Disease Assessment and Mass Spectrometry

MRD was assessed by the next-generation sequencing (NGS) ClonoSEQ® assay (Adaptive Biotechnologies, Seattle, WA, USA) on bone marrow samples from patients who achieved at least a very good partial response (VGPR) at cycle 9 (C9) or later. When BM was available, a second MRD assessment was performed to confirm the MRD result. Quality control (QC) of NGS results was performed as previously described^37^.

Minimal residual disease by next-generation sequencing was evaluable in 25 patients who achieved at least a VGPR. Baseline pre-treatment bone marrow samples were available for 22/25 patients and were analyzed for the identification of the VDJ molecular marker by the Adaptive MRD assay. Twenty out of 22 (91%) baseline samples passed the QC analysis, and the molecular marker was identified in 18/22 patients (82%). For MRD analysis of these 18 patients, bone marrow samples that were available from end of induction (C9) and from the end of treatment (EOT) were used for sequencing. Three of the 18 patients did not have any sample available for MRD analysis and one sample had indeterminate results at limits of detection, 13 patients had MRD testing at the end of induction (C9), 10 at EOT, and 8 individuals had an assessment at both timepoints for an overall total of 23 samples tested for MRD.

Residual disease after therapy was also assessed by matrix-assisted laser desorption ionization-time of flight (MALDI-TOF) mass spectrometry to quantify M proteins (Binding Site Group, Birmingham, UK), and Optilite free light-chain assay to quantify IgG, IgA, IgM, and serum free light chains. EXENT-iQ software (Binding Site Group, Birmingham, UK) was used for quantitative analysis of detected M proteins. The lower limit of accurate M protein quantification by MALDI-TOF mass spectrometry was 0·015 g/L. Serum (500 μL) was tested using the EXENT system and immunoglobulin (Ig) isotype assay^38^. The unique mass/charge (m/z) (+/− 5) of a monoclonal light chain and the isotype were defined at baseline and tracked in each patient’s samples after therapy at the end of induction (C9) and when available during maintenance (2-3 months after C9) and at end of treatment (EOT). MALDI-TOF was performed at baseline before treatment in 48 patients out of 55.

Seven out of 55 patients were excluded due to sample unavailability (n = 5/7) and light chain-only disease at baseline (n = 2 out of 55), defined as having a negative SPEP and serum immunofixation (IFX) result in the setting of SMM (**Suppl Figure S2**). We assessed the concordance between Isotype by MALDI-TOF and SPEP-IFX and the correlation between quantified M protein at baseline with the two methods using Pearson correlation. The concordance between NGS MRD results and residual disease by MALDI-TOF was also assessed by comparing the presence of residual M protein >0·015 g/L (limit of detection)^39^ and positivity at 10^−5^ by NGS. Concordance between MS by MALDI-TOF, IFX, and NGS in BM was assessed using Cohen’s kappa (κ) statistic, with κ value interpretation per Landis and Koch^40^.

### DNA isolation and library construction

Genomic DNA isolation was carried out using the Monarch Genomic DNA Purification Kit (New England Biolabs), with tumor (PCs) and normal (PBMCs) yields quantified by Qubit 3.0 fluorometer (Thermo Fisher Scientific). 50-100ng was taken forward for DNA sequencing library preparation using the NEBNext Ultra II FS DNA Library Prep kit (New England Biolabs) with unique dual index adapters (NEBNext Multiplex Oligos) according to manufacturers’ instructions. Final library fragment sizes were assessed using the BioAnalyzer 2100 (Agilent Technologies), and yields were quantified by Qubit 3.0 fluorometer (Thermo Fisher Scientific) and qPCR (KAPA Library Quantification Kit).

### Whole genome sequencing (WGS) and genomic data analysis

WGS was available for 17 patients. Final sample libraries were normalized and pooled before WGS was performed on Illumina NovaSeq 6000 S4 flow cells, 300 cycles 2×150bp paired-end reads, at the Genomics Platform of the Broad Institute of MIT and Harvard. WGS analysis was performed on an in-house cloud-based HPC system for copy-number and structural variant analyses, as previously described^41^. Sequencing reads were aligned to the GRCh38 reference genome with the bwa mem v0.7.7 algorithm^42^, duplicate reads were marked with MarkDuplicates from picard v1.457, indels were realigned with GATK 3.4 IndelRealigner, and base qualities were recalibrated with GATKBQSR. WGS analysis was performed with the Cancer Genome Analysis workflow from the Cancer Program at Broad Institute of MIT and Harvard on an in-house cloud-based HPC system. Small indels were called with Strelka2 and were filtered (i) against a panel of normals (PoN), (ii) for potential technical artifacts (oxoG), and (iii) for multiple alignment with BLAT^43^. After copy-number normalization with AllelicCapSeg, ABSOLUTE solutions were manually reviewed to estimate mutation CCF, purity, and ploidy of tumor samples^44^. Structural variants were detected and filtered as previously described^45^.

### Patient sample processing for scRNA-seq

Whole bone marrow aspirates (5-20mLs) were drawn into EDTA preservative tubes and kept at 4°C before being processed within a 6 hours from collection. Bone marrow mononuclear cells were isolated from aspirates using density gradient centrifugation. Briefly, the bone marrow was filtered to remove any clots or bone debris, diluted 1:3 with 1X Phosphate-Buffered Saline (PBS), and gently poured above 15mLs of Ficoll-Paque density gradient medium within a SepMate™ PBMC isolation tube (StemCell Technologies, Cat # 85450). After centrifugation at 1200 rcf/g for 15 minutes, PBMCs were poured away from unwanted cells and washed twice with ice-cold 1xPBS prior to plasma cell enrichment. Plasma cells were enriched from the bone marrow samples using CD138 magnetic bead separation using single-column positive selection (Miltenyi Biotec, Cat #130-097-614).

### Single-cell RNA/V(D)J library construction and sequencing

Sequencing libraries were prepared from BM samples, including 5’ scRNA-seq libraries and scBCR-seq and scTCR-seq libraries. After MACS enrichment, all CD138+ and CD138-cells were centrifuged at 300 rcf/g for 5 mins and washed twice with an ice-cold 0.1% Ultrapure Bovine Serum Albumin (BSA)/PBS wash buffer. Subsequently, cells were either subjected to volume reduction or diluted based on cell counts obtained using both a hemocytometer and a Countess automated cell counter (ThermoFisher) and cell mixtures were adjusted to obtain optimal cell densities for achieving maximal cell recovery. Samples, Gel bead-in-EMulsion (GEMs), and partitioning oil were loaded into a Next GEM Chip K microfluidic device and placed in a Chromium Controller instrument for downstream single-cell encapsulation and recovery. All GEM generation/barcoding, post GEM RT clean-up/cDNA amplification and 5’ gene expression (GEX) library construction steps were completed using the Chromium Next GEM Single Cell 5’ Reagent Kit v2 (Dual Index) and Library Construction Kit, according to the manufacturer’s instructions. cDNA generated was also subsequently subjected to V(D)J amplification using Chromium Single Cell Human BCR Amplification Kit and Library Construction Kit for scBCR-seq and Chromium Single Cell Human TCR Amplification Kit and Library Construction Kit for scTCR-seq, according to the manufacturer’s instructions. The quality of sample libraries was assessed based on library trace fragment sizes and patterns at numerous points throughout the protocol including cDNA generation, BCR V(D)J pre-amplified cDNA generation, TCR V(D)J pre-amplified cDNA generation, final GEX, final BCR and final TCR construction steps using a High-Sensitivity DNA Analysis Kit and the Bioanalyzer 2100 instrument (Agilent Technologies). Final GEX, BCR and TCR library quantification was performed using Quant-iT Picogreen dsDNA Assay Kit (Invitrogen) before preparing single pools for sequencing. Pooled libraries were sequenced on NovaSeq S4 flow cells at the Genomics Platform of the Broad Institute of MIT and Harvard (Cambridge, MA).

### Single-cell RNA/V(D)J data processing

CD138+ scRNA-seq was available for 13 patients, CD138-scRNA-seq was available for 20 patients and 12 patients had paired CD138+ and CD138-scRNA-seq. Paired WGS and CD138+ RNA sequencing was available for 10 patients. 20 patients had either scRNA-seq or WGS performed on their tumor samples.

CellRanger mkfastq (v5.0.1) was used to generate FASTQ files^46^. Gene expression matrices were generated by CellRanger count (v6.0.1) with the genome reference (refdata-gex-GRCh38-2020-A) provided by 10X Genomics^46^. To remove ambient RNA, CellBender (v0.2.0) was run on the gene expression matrices with the target false positive rate cutoff of 0.01. Poor quality cells with >15% mitochondrial gene expression, either <200 detected genes, >5,000 detected genes, <400 UMIs, or >50,000 UMIs were filtered out. Three doublet tools, Scrublet (v0.2.3), scDblFinder (v1.8.0), and scds (1.10.0) were used to calculate multiplet scores^47–49^. CellRanger vdj (v6.0.1) was used on FASTQ files with the VDJ reference file (refdata-cellranger-vdj-GRCh38-alts-ensembl-5.0.0)^46^.

### Malignant plasma cell identification

Plasma cells were first identified based on expressing key lineage markers, such as *SDC1* (encoding CD138), *CD38*, *XBP1*, *PRDM1*, *IRF4*, and *TNFRSF17* (encoding BCMA). Cell barcodes that were determined to correspond to plasma cells were considered for downstream analysis. Tumor cells were identified on the basis of belonging to the largest expanded BCR clone and clustering separately from normal plasma cells. For all clones, the isotype of the malignant clone matched that detected clinically via IFX. We annotated 50,591 of patient cells as tumor, and the rest (n=1876) as healthy plasma cells based on our tumor cell annotation protocol. The median number of tumor cells per patient was 2,148.

### Single-cell TCR sequencing data processing and analysis

We performed single-cell TCR sequencing on CD138-samples with available single-cell RNA sequencing data (n=9). Complementary DNA generated from barcoded CD138-immune cells using Chromium Single Cell 5’ Gene Expression and V(D)J enrichment kits by 10X Genomics was subjected to V(D)J enrichment and library preparation and sequenced on a NovaSeq 6000 instrument at the Genomics Platform of the Broad Institute of MIT & Harvard.

CellRanger v5.0.1 was used to extract FASTQ files and produce clonotype matrices^50^. When multiple alignments were called for a single chain, the alignment with the most UMIs was selected, and when multiple chains were called for a single cell barcode, the top two chains in terms of UMI counts were selected.

Clone size assignment:

To categorize TCR clones based on its size or its abundance within a sample, we employed a downsampling approach to account for potential sampling bias. We randomly sampled 100 T cells from each patient sample 100 times. Within each iteration, we calculated the proportion of each unique TCR clone compared to the total T cells sampled. Finally, for each clone, we averaged its proportional abundance across all iterations to obtain a more stable estimate of its size category. Categories were defined as: Rare: ≤1%, Small: > 1% and < 5%, Medium: ≥ 5% and < 10%, Large: ≥ 10%.

Clone-size-category proportion estimation within cell subtypes per patient sample:

To estimate the overall proportion of clones belonging to each size category within a cell subtype per patient sample, we again used downsampling. We randomly sampled 100 T cells from each patient cell subtype sample 100 times. In each iteration, we counted the frequency of each clone size category within the downsampled set. Finally, we averaged the frequency of each category across all iterations and these averaged frequencies were then renormalized before plotting. Two-sided p-values were computed by bootstrapping with 1000 iterations.

### Copy number analysis from scRNA sequencing

Copy number abnormalities on tumor cells were inferred using Numbat with default parameters (v1.1.0)^22^. Allelic data was collected from plasma cells and B cells and a panel of 1,200 plasma cells from 11 healthy donors was used as expression reference.

### Differential Expression Analysis

Droplets with < 20,000 reads per cell were discarded, and 20,000 reads per cell were downsampled across the entire CD138pos scRNA-seq cohort. To control the influence of individual tumors on the analysis, as well as control for the lower number of non-progressors in this sub-cohort (n=3, compared to 10 progressors), we randomly sampled the same number of tumor cells from progressors and non-progressors (∼2100 per group) and balanced the contribution of each individual towards the group (progressors: 210 cells per patient as one progressor patient had fewer than 210 cells (n=72) after downsampling; non-progressors: 700 cells per patient). We repeated this process leaving out the one progressor patient with low number of cells this time considering only 9 progressor patients and 3 non-progressors and once again balancing the contribution of each individual towards the group (progressors n=225, non-progressors n=675 per patient). No difference in results was observed.

### Statistical Analysis

The primary outcome was progression-free survival after two years of study treatment. Progression-free was defined as being followed for at least two years from the start of protocol therapy and no confirmed disease progression. The primary endpoint was calculated for both progression to SLIM-CRB criteria as well as biochemical progression.

For this manuscript, progressive disease was defined as per the IMWG uniform response criteria for MM with either SLIM-CRAB progression or a rise of 25% in serum or urine M protein or difference between involved and uninvolved FLC levels^51^.

Patients who did not have at least 2 years of follow-up or progressed between the start of treatment and two years after EOT were counted as failed progression-free at 2 years. The study was designed for a 50 vs 70% progression-free rate with 5% and 10% type I and type II errors, respectively. The critical value to reject the null hypothesis of 50% was 33 or more of 53 patients and was evaluated using a one-sided exact binomial test.

We report the primary outcome and responses to study treatment as proportions with 90% exact binomial confidence intervals. Continuous measures are summarized by median and range, and categorical variables are summarized as proportions.

Categorical variables were tested for association with continuous and other categorical variables using Wilcoxon rank-sum (or Kruskal-Wallis for three or more groups) or Fisher’s exact tests, respectively. Time-to-event endpoints are estimated using the method of Kaplan and Meier with 95% confidence intervals calculated using Greenwood’s method to estimate variance. Median follow-up is calculated using the reverse Kaplan-Meier method. Statistical analyses were performed using R version 4.1.2 (2021-11-01).

## Acknowledgments

The authors would like to thank the patients and their families for participating in this study.

Anna V. Justis, PhD, a medical writer employed by Dana-Farber Cancer Institute, edited this manuscript under the direction of the authors.

## Authorship Contributions

O.N.: Writing-Original Draft, Writing-Review & Editing

M.P.A.: Writing-Original Draft, Investigation, Formal analysis, Visualization, Writing-Review & Editing

R.A.R.: Formal analysis, Visualization, Writing-Review & Editing

M.T: Formal analysis, Visualization, Writing-Review & Editing

S.M.: Writing-Review & Editing

J.A. Writing-Review & Editing, Supervision

L.B.: Investigation, Writing-Review & Editing

A.D. Writing-Review & Editing, Supervision

H.E.: Investigation, Writing-Review & Editing

M.B.: Investigation, Writing-Review & Editing

E.D.L.: Investigation, Writing-Review & Editing

J.P.L.: Writing-Review & Editing

G.B.: Writing-Review & Editing

E.O.: Writing-Review & Editing

T.W.: Writing-Review & Editing

J.T.: Writing-Review & Editing

K.A.: Writing-Review & Editing

P.G.R.: Writing-Review & Editing

G.G.: Writing-Review & Editing

L.T.: Writing-Review & Editing, Supervision

R.S.P.: Writing-Review & Editing, Supervision

I.M.G.: Conceptualization, Resources, Writing-Review & Editing, Supervision, Funding Acquisition

## Inclusion and Ethics

One or more of the authors of this paper self-identifies as an underrepresented ethnic and/or gender minority in science. One or more of the authors of this paper self-identifies as a member of the LGBTQIA+ community. We support inclusive, diverse and equitable conduct of research.

## Data availability

Single-cell RNA and TCR-sequencing raw data generated for this study will be deposited in dbGaP (study site pending). Gene expression matrices can be accessed on Mendeley: https://data.mendeley.com/preview/z56k3y8cdg?a=6945f72a-b190-4fb1-b0fe-31c12a70a0d4 (reserved DOI:10.17632/z56k3y8cdg.1)

## Code Availability

This paper does not report original code.

