## Supplemental Figures for "Long-Term Follow-Up Defines the Population That Benefits from Early Interception in a High-Risk Smoldering Multiple Myeloma Clinical Trial Using the Combination of Ixazomib, Lenalidomide, and Dexamethasone"

### Slide 1
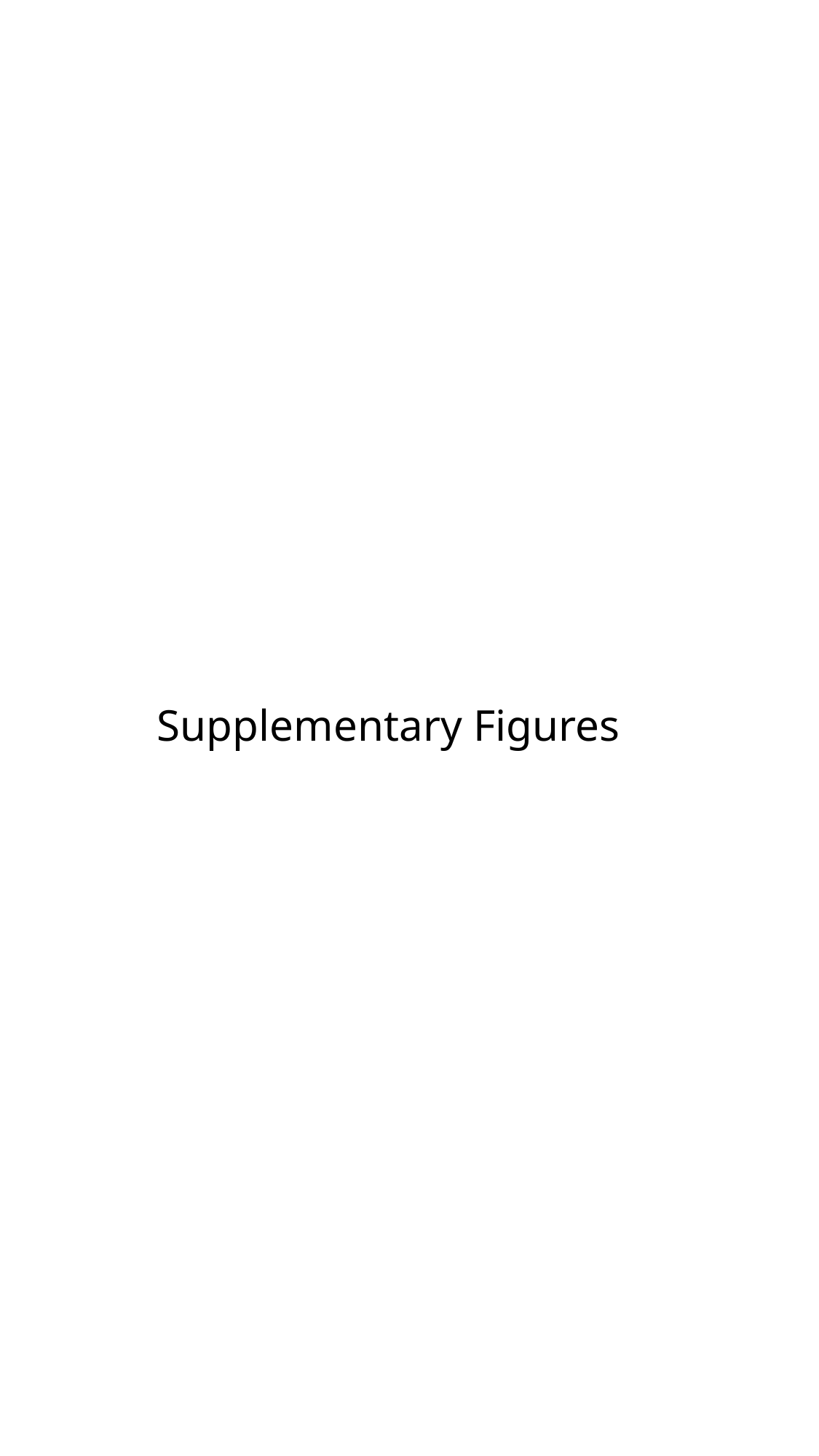

#### Supplementary Figures

### Slide 2
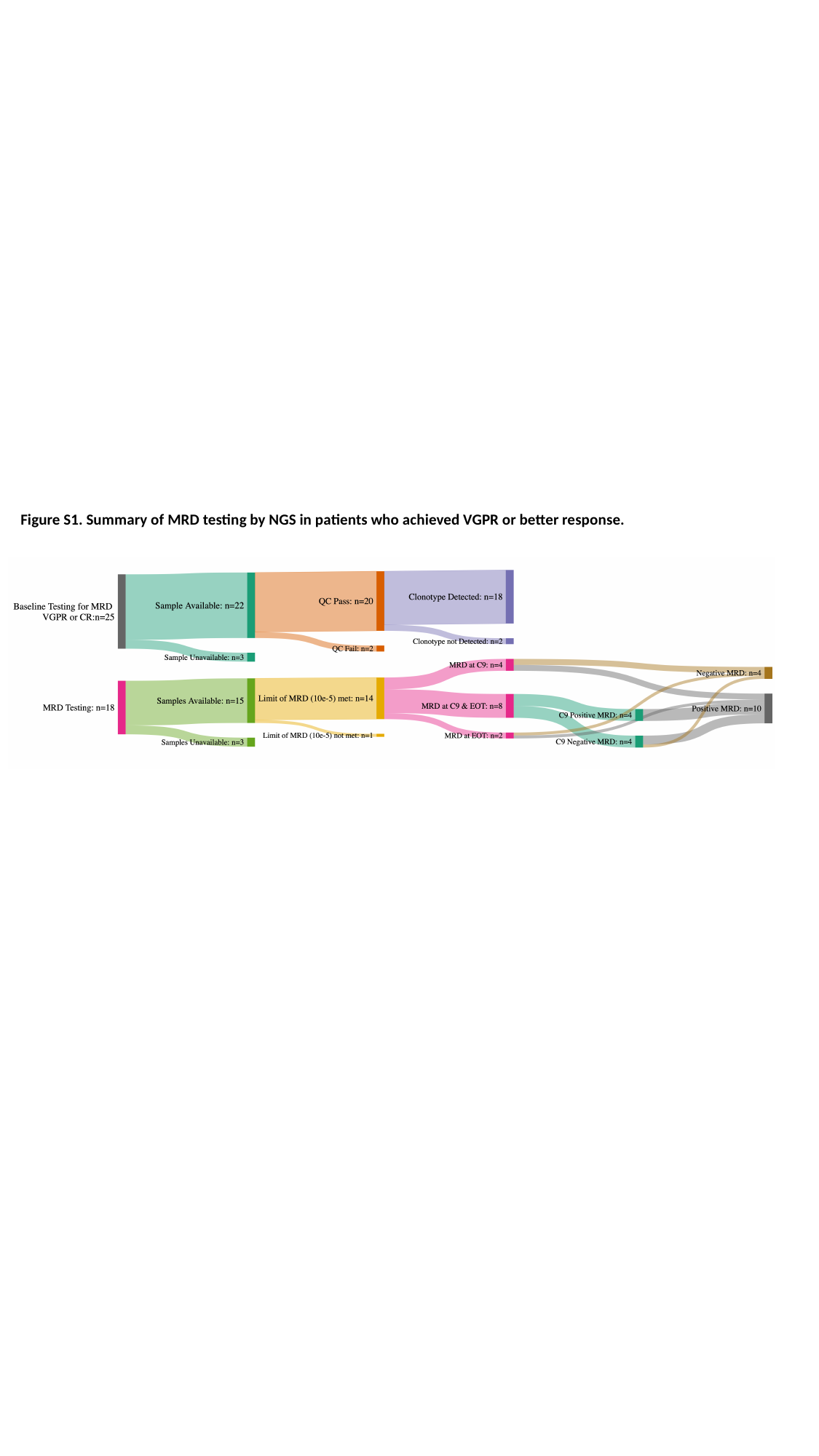

Figure S1. Summary of MRD testing by NGS in patients who achieved VGPR or better response.

### Slide 3
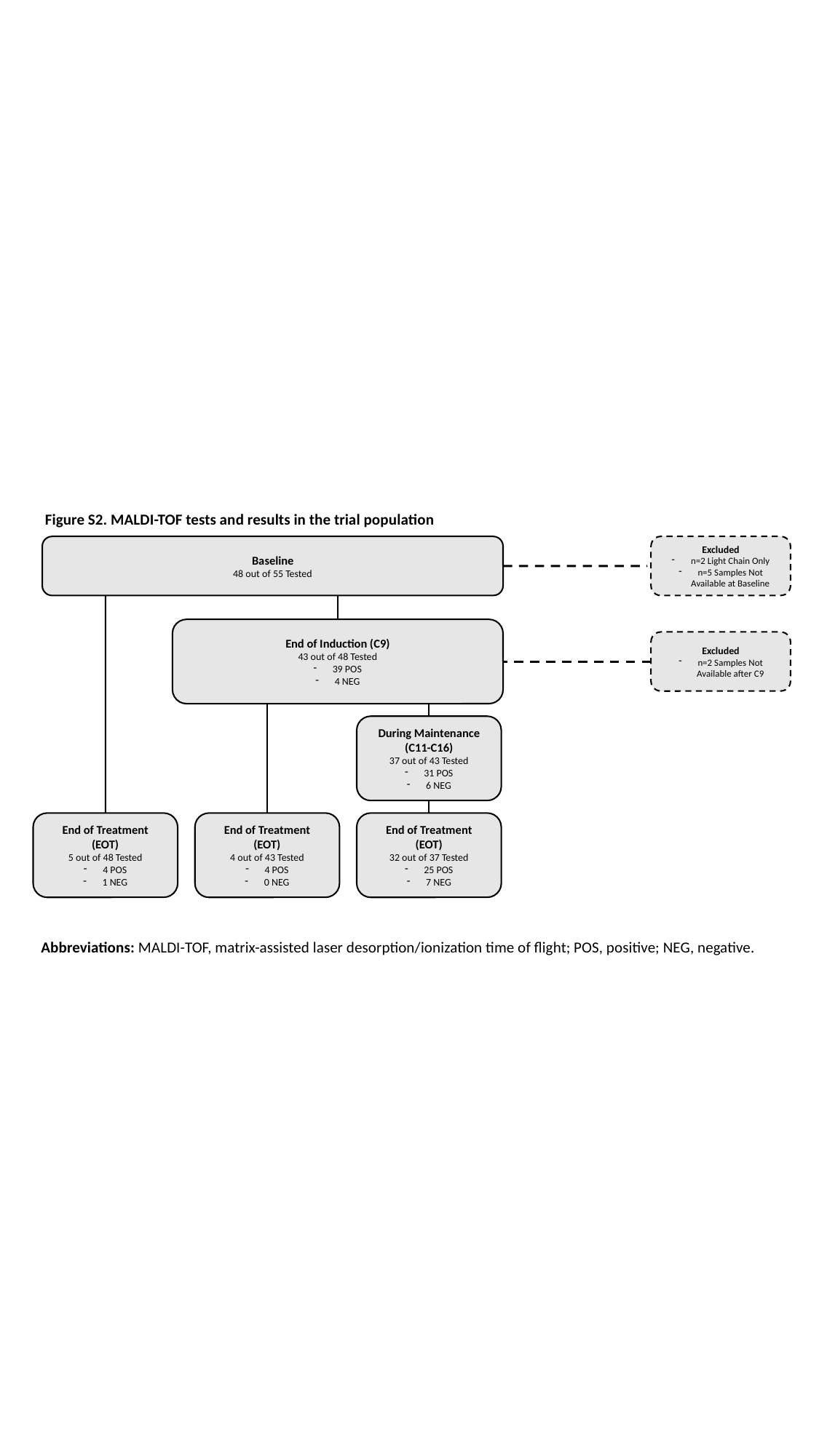

Figure S2. MALDI-TOF tests and results in the trial population​
Baseline
48 out of 55 Tested
Excluded
n=2 Light Chain Only
n=5 Samples Not Available at Baseline
End of Induction (C9)
43 out of 48 Tested
39 POS
4 NEG
Excluded
n=2 Samples Not Available after C9
During Maintenance (C11-C16)
37 out of 43 Tested
31 POS
6 NEG
End of Treatment (EOT)
5 out of 48 Tested
4 POS
1 NEG
End of Treatment (EOT)
4 out of 43 Tested
4 POS
0 NEG
End of Treatment (EOT)
32 out of 37 Tested
25 POS
7 NEG
Abbreviations: MALDI-TOF, matrix-assisted laser desorption/ionization time of flight; POS, positive; NEG, negative.​

### Slide 4
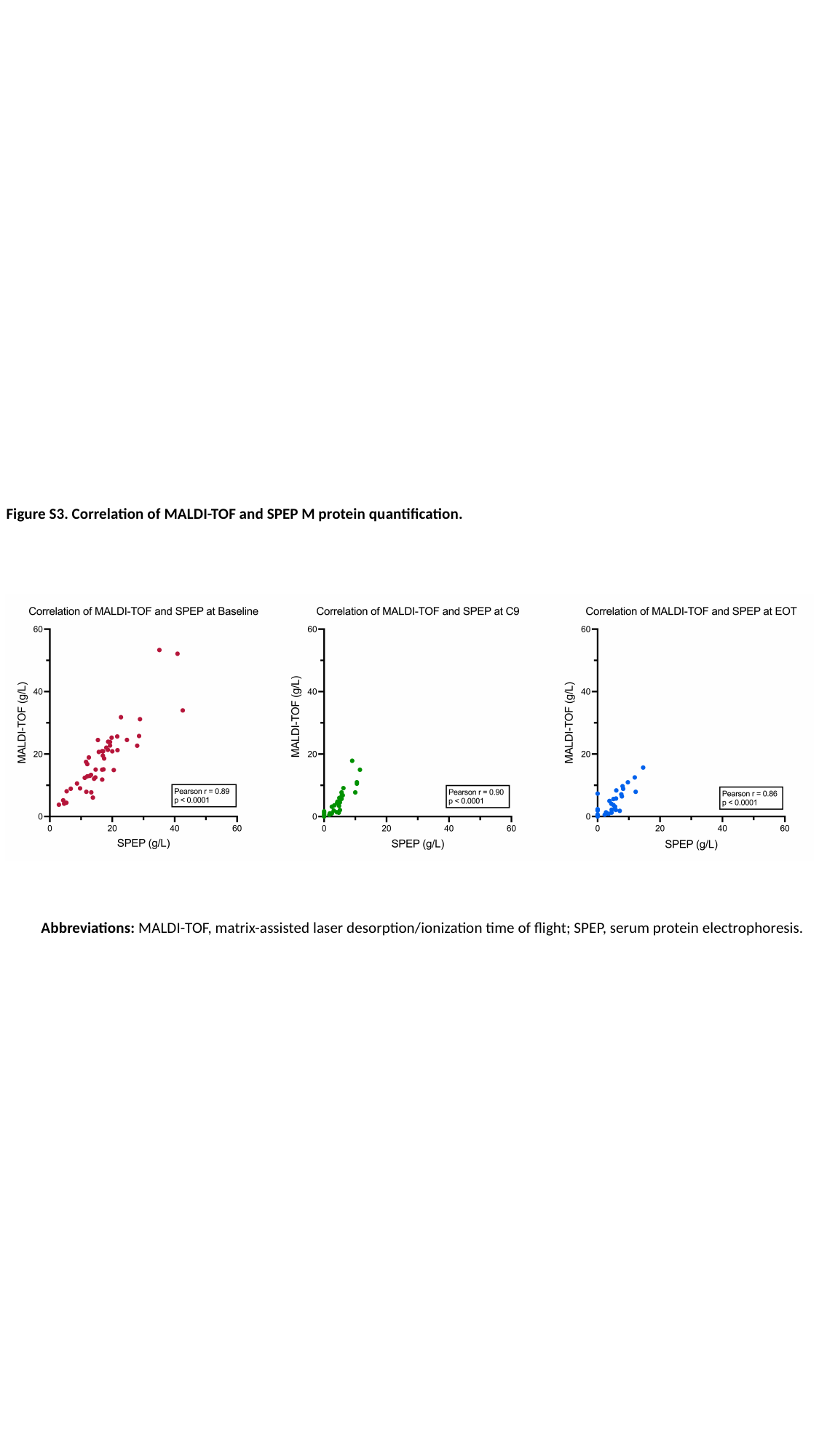

Figure S3. Correlation of MALDI-TOF and SPEP M protein quantification.
Abbreviations: MALDI-TOF, matrix-assisted laser desorption/ionization time of flight; SPEP, serum protein electrophoresis.

### Slide 5
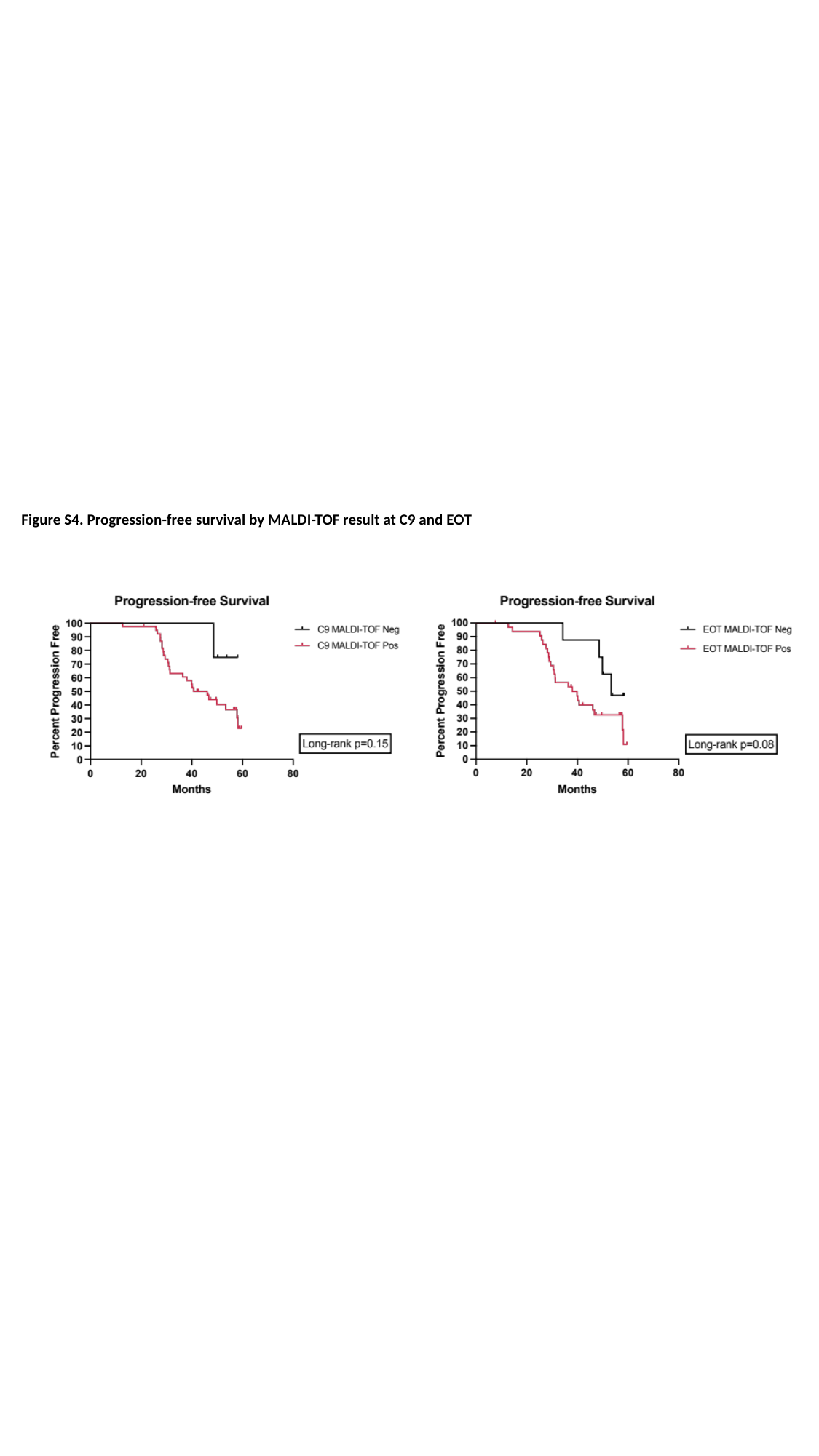

Figure S4. Progression-free survival by MALDI-TOF result at C9 and EOT
