## Supplemental Tables for "Long-Term Follow-Up Defines the Population That Benefits from Early Interception in a High-Risk Smoldering Multiple Myeloma Clinical Trial Using the Combination of Ixazomib, Lenalidomide, and Dexamethasone"

**Table S1. Concordance of MALDI-TOF and IFX results after therapy.**

| **Test Result** | | **End of Cycle 9** | **End of treatment** | **All cases** |
| --- | --- | --- | --- | --- |
| **IFX** | **MALDI-TOF** |  |  |  |
| (+) | (+) | 36 | 28 | 64 |
| (-) | (-) | 4 | 8 | 12 |
| (+) | (-) | 0 | 0 | 0 |
| (-) | (+) | 4 | 6 | 10 |
| Kappa Statistic  (Cohen’s κ, 95%CI) | | 0.69 [0.29, 0.95] | 0.64 [0.39, 0.89] | 0.64 [0.44, 0.84] |
| Percent agreement | | 91% | 86% | 88% |
| PPV | | 90% | 82% | 86% |
| NPV | | 100% | 100% | 100% |

**Abbreviations:** MALDI-TOF, matrix-assisted laser desorption/ionization time of flight; IFX, serum immunofixation; PPV, positive predictive value; NPV, negative predictive value
